# Optimal mix of differentiated service delivery models for HIV treatment in Zambia: a mathematical modelling study

**DOI:** 10.1101/2024.06.17.24309039

**Authors:** Nkgomeleng Lekodeba, Sydney Rosen, Bevis Phiri, Sithabiso Masuku, Caroline Govathson, Aniset Kamanga, Prudence Haimbe, Hilda Shakwelele, Muya Mwansa, Priscilla Lumano-Mulenga, Amy Huber, Sophie Pascoe, Lise Jamieson, Brooke E Nichols

## Abstract

**Background:** Zambia has scaled up differentiated service delivery (DSD) models for antiretroviral treatment (ART) to provide more client-centric care and increase service delivery efficiency. The current DSD landscape includes multiple models of care based on guidelines, resources, partner inputs, and other factors. We used local data to identify cost-effective combinations of DSD models that will maximize benefits and/or minimize costs to guide future DSD expansion.

**Methods:** We developed a mathematical Excel-based model using retrospective retention and viral suppression data from a national cohort of ART clients (≥15 years) between January 2018-March 2022 stratified by age, sex, setting (urban/rural), and model of ART delivery. Outcomes (viral suppression and retention in care), provider costs, and costs to clients for each model were estimated from the cohort and previously-published data. For different combinations of the nine DSD models in use, we evaluated the incremental cost to the health system per additional ART client virally suppressed on treatment compared to the 2022 base case.

**Results:** Of the 125 combinations of DSD models evaluated, six were on the cost-effectiveness frontier (CEF): 1) six-month dispensing (6MMD)-only; 2) 6MMD and adherence groups (AGs); 3) AGs-only; 4) fast track refills (FTRs) and AGs; 5) FTRs-only; and 6) AGs and home ART delivery. 6MMD-only was cost-saving compared to the base case, increased the proportion of clients virally suppressed by 1.6%, and decreased costs to clients by 16.6%. The next two scenarios on the CEF, 6MMD+AGs and AGs-only, each cost an additional $245 per person virally suppressed, increased the total number of individuals suppressed on treatment by 2.8% and 4.8%, respectively, and increased costs to clients by 63% and 143%, respectively.

**Conclusions:** Mathematical modelling using existing data can identify cost-effective mixes of DSD models and allocations of clients to these models, while ensuring that all client sub-populations are explicitly considered. In Zambia, providing 6MMD to all eligible clients is likely to be cost-saving, while health outcomes can be improved by allocating clients to selected models based on sub-population.

## Introduction

Many countries in sub-Saharan Africa are expanding differentiated service delivery (DSD) for HIV treatment to efficiently achieve national and global targets for treatment uptake and viral suppression [1,2]. DSD models for HIV treatment differ from conventional care in several ways: the location of service delivery, frequency of interactions with the healthcare system, types of services provided, and cadres of staff involved. They are designed to provide a more client-centred approach, aiming to improve outcomes of those on antiretroviral treatment (ART) and reducing costs to and improving the experiences of both clients and healthcare providers [1,2]. The World Health Organization has recommended the implementation of differentiated service delivery for HIV treatment since 2015 [3], and many countries, including Zambia, have incorporated DSD models into national HIV treatment guidelines [4].

In Zambia, a high HIV-prevalence country with an estimated 1.4 million people living with HIV [3], the Ministry of Health currently supports a range of “low intensity” DSD models that require fewer resources from both clients and providers than does conventional ART care [5]. Models described in national guidelines include 6-month dispensing of ART at facilities (6MMD), community medication pickup points, appointment spacing, fast-track dispensing at facilities, and other models targeted specific population groups, such as teen/scholar and extended clinic hours models. Conventional care typically entails 3-month dispensing of ART at health facilities, with a clinical consultation and medication pickup at each quarterly clinic visit. Each public sector HIV clinic offers some combination of these models, based on available resources and staff choices. In addition, non-governmental partner organizations support several other bespoke models at subsets of healthcare facilities.

As Zambia continues to improve differentiated service delivery to build the long-term sustainability of its HIV program, more attention is needed to the optimal combination of models that facilities should offer. Ideally, the allocation of models should reflect considerations of the trade-offs and synergies among health outcomes, costs and benefits to clients, and costs and benefits to the healthcare system. While some evidence is available about each of these factors [2,6–10], little is known about the cost-effectiveness of different combinations of DSD models, at any level. In most settings, in Zambia and elsewhere, the distribution of models across facilities and nationally is largely determined by national guidelines, which themselves are generally not tailored to local conditions, and by available facility and partner resources.

To assist in the prioritization and scale-up of cost-effective DSD models, we developed the Alternative Delivery of ART Optimization (ADAPT) model, an Excel-based mathematical model that provides a decision-making framework for scaling-up differentiated service delivery for ART. ADAPT compares different DSD model implementation scenarios in terms of viral suppression and providers’ and clients’ costs at a national level. We parameterised ADAPT for Zambia, which has a rich endowment of both routine and research data sources, and then conducted a cost-effectiveness analysis of likely combinations of ART delivery models to demonstrate how model mix and allocation may affect achievement of HIV program goals.

## Methods

We developed and parameterised an Excel-based mathematical model of all ART clients in care in Zambia using de-identified, retrospective SmartCare electronic data from Zambia’s national cohort of ART clients (aged ≥15 years) from January 2018 to March 2022 (Table 3) [11]. SmartCare, Zambia’s public sector electronic medical record system for HIV, captures roughly 75% of all ART patients in Zambia [12,13]. It includes data fields indicating when an ART client enrolled in a DSD model, the specific model enrolled in, the number of months for which ART medications were dispensed at each event, treatment outcome, outcome date, and demographic characteristics [12,14].

### Model structure and approach

To determine the expected impact and cost-effectiveness of the implementation of various combinations of DSD models and uptake in each model, we started with a base case representing the actual 2022 national distribution of ART clients among DSD models disaggregated by population sub-groups (age group, sex, urban or rural location). The health outcomes from this base case reflect the health outcomes observed in 2022 for all DSD models for each population sub-group.

We then constructed multiple hypothetical scenarios in which specific combinations of one or more DSD models can either be implemented across the entire population cohort or be targeted to specific sub-populations based on age, sex, and urban or rural setting. Each scenario reflected a specific distribution of ART clients among different models. The model then generated a total number of individuals virally suppressed and retained in care (sum of all individuals virally suppressed on treatment and retained in care by model of ART delivery for each scenario), and a total estimated cost to the provider and clients. The modelling approach, comprised of five distinct steps, is depicted in Figure 1.

**Figure 1.**
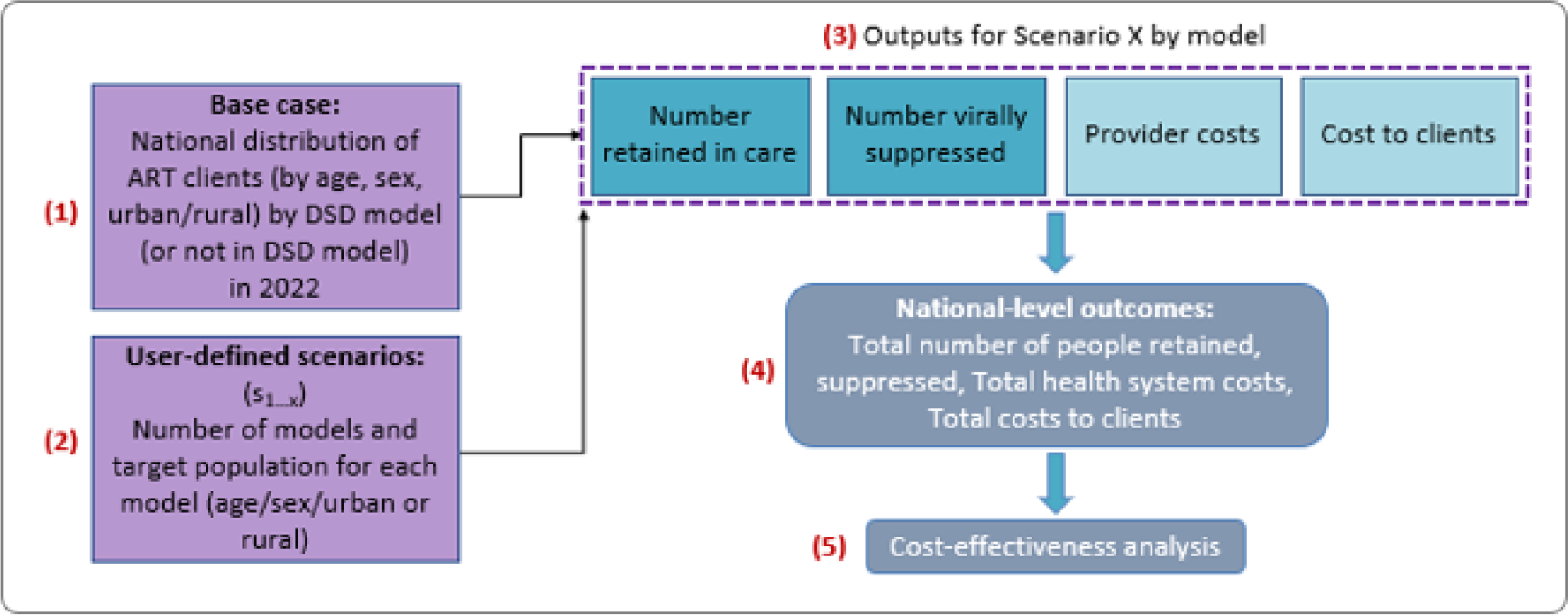
ADAPT modelling structure

### Cohort, eligibility and outcomes

The primary outcomes for this analysis were retention in care, viral suppression, provider costs, and cost to ART clients. Viral suppression and provider costs were combined into an incremental cost-effectiveness ratio for each model. Study cohort, eligibility criteria and outcomes are defined in Table 1.

**Table 1.** Definitions of study cohort, eligibility criteria and outcomes.

| Cohort, criterion, or outcome | Definition |
| --- | --- |
| *Study cohort* |  |
| Study analytic cohort | The study analytical cohort comprised ART clients (aged $\geq 15$ years) included in the SmartCare database in January 2018 to March 2022 and enrolled in differentiated or conventional care. |
| *Criteria* |  |
| Eligibility for DSD enrollment | On ART for at least 12 months. (Because there were limited viral load data available, viral suppression could not be used as an eligibility criterion despite it being specified in DSD policy guidelines) |
| DSD model assignment | Each participant was assigned to the first DSD model they enrolled in during the first year after becoming eligible for DSD. Those who did not have any DSD model enrollment reported in the dataset were classified as remaining in conventional care |
| *Outcomes* |  |
| Total health system costs | The sum of provider costs to the health system per year |
| Total costs to clients | The sum of cost to clients incurred accessing ART services per year |
| Retention in care at 12 months | The proportion of ART clients who had a clinic visit between 11 and 24 months stratified by age group, sex, setting (urban/rural), and model of ART delivery. Retention in care for clients enrolled in DSD model was estimated after they became eligible and were enrolled in DSD models. |
| Viral suppression at 12 months | The proportion of ART clients who had viral load test results of <1,000 copies/ml at 12 months, stratified by age group, sex, setting (urban/rural), and model of ART delivery. The viral load at 12 months is measured between 11 and 24 months. |
| Incremental cost-effectiveness ratio (ICER) | Healthcare system (provider) cost per additional ART client virally suppressed on treatment |

### ART delivery models

In the modelled scenarios we considered a total of 11 models of ART delivery for ART clients in 2022 (Table 2). Two were variations of conventional (non-differentiated) care: 3-month facility-based dispensing (3MMD), which is the current standard of care, and a study-defined model for clients observed to be receiving less than 3 months of medication at a time. Nine were DSD models [8,15]. At the time of the study, Zambia’s criteria for being designated as “established on treatment” were a viral load of <1,000 copies/mL and a minimum of 6 months on ART. ART clients in conventional care went through all service points (registration, vitals, clinical consultation, pharmacy, etc.) within the facility at each quarterly visit.

**Table 2.**
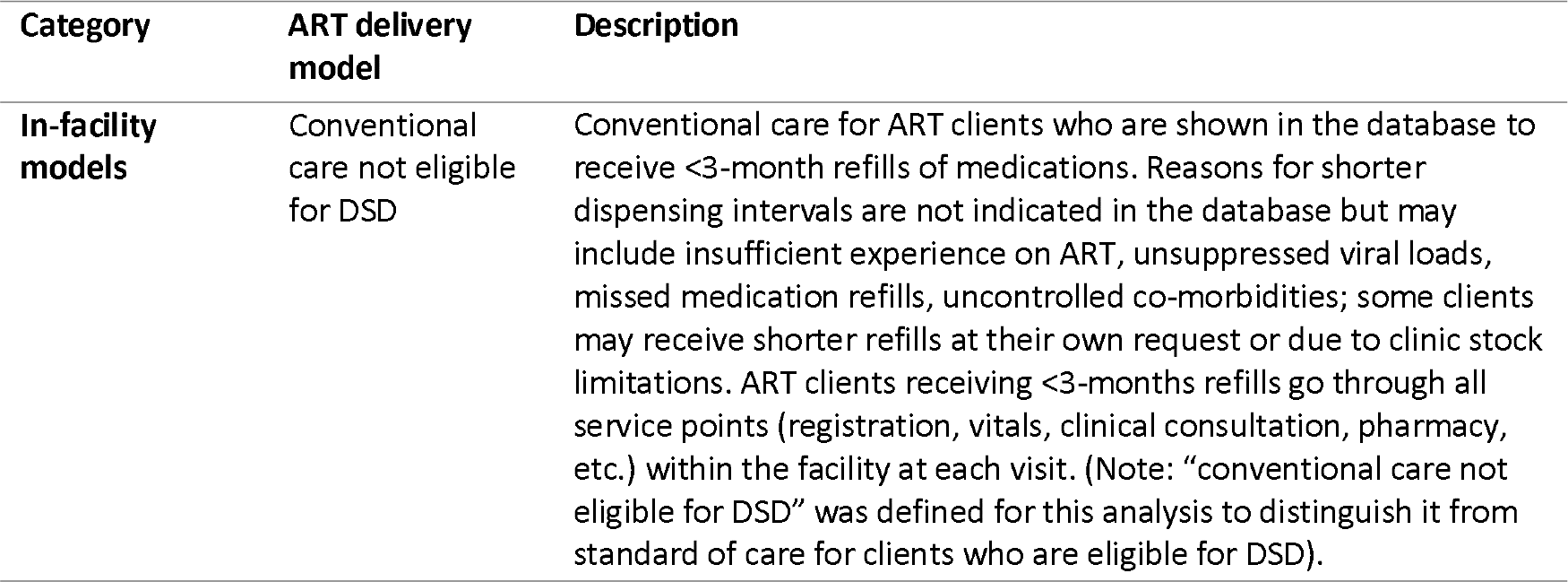
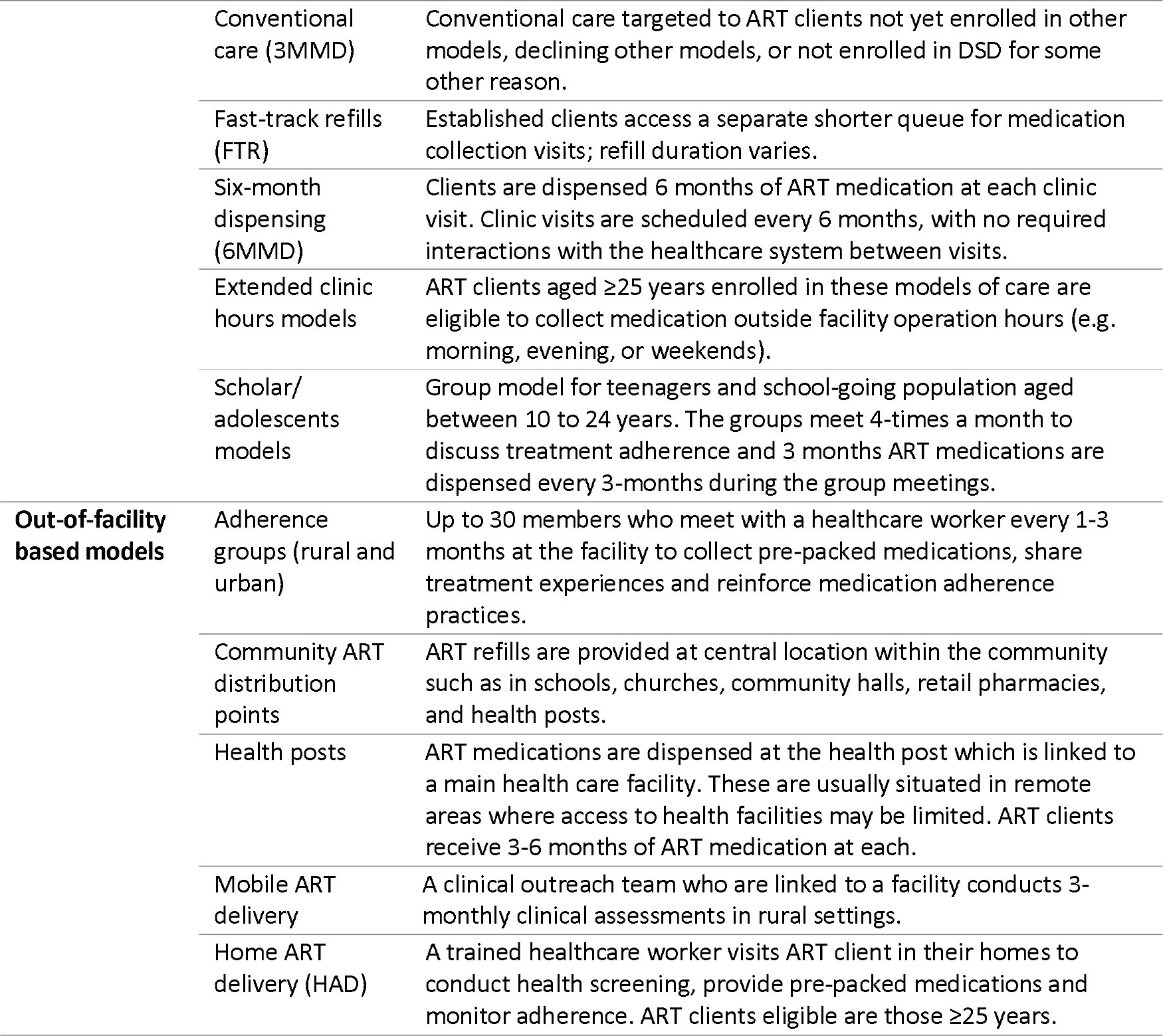
Description of conventional and differentiated ART service delivery models.

The nine DSD models considered in the analysis were categorised as either in-facility or out-of-facility models and are described in detail in Table 2 [4]. We did not include models of care tailored to clients with viral loads >1,000 copies/ml, such as “high viral load clinics.” Instead, cohort participants who did not meet DSD eligibility criteria were retained in conventional care in our mathematical model.

### Input parameters and assumptions

Model input parameters, parameter values, client characteristics, and the base case distribution of clients that reflect the 2022 cohort are presented in Table 3. Health outcomes (retention in care and suppression rates) for each ART delivery model, stratified by sex, age, and setting, were estimated from the SmartCare database (Table 3, Table S1) [10].

**Table 3.** Baseline input parameters (N=917,749)

| Parameter | Model value, n (%) |  |
| --- | --- | --- |
| *Demographic characteristics* [11] |  |  |
| Sex (female) | 584,509 (63.7%) |  |
| Setting (urban) | 636,597 (69.3%) |  |
| Age group |  |  |
| 15-19 years | 21,608 (2.4%) |  |
| 20-24 years | 49,990 (5.4%) |  |
| 25-49 years | 660,085 (71.9%) |  |
| 50+ years | 186,066 (20.3%) |  |
| *Distribution of ART delivery models (n, %) [11]* | *Male* | *Female* |
| Conventional care not eligible for DSD | 17,914 (5.4%) | 30,766 (5.3%) |
| Conventional care (3MMD) | 47,302 (14.2%) | 85,728 (14.7%) |

| Parameter | Model value, n (%) |  |  |  |
| --- | --- | --- | --- | --- |
| Six months dispensing | 231,153 (69.4%) | 401,085 (68.6%) |  |  |
| Fast-track refills | 29,096 (8.7%) | 52,691 (9.0%) |  |  |
| Extended clinic hours | 75 (<0.1%) | 130 (<0.1%) |  |  |
| Scholar/adolescent model | 92 (<0.1%) | 110 (<0.1%) |  |  |
| Mobile ART distribution | 583 (0.2%) | 1,159 (0.2%) |  |  |
| Health post | 691 (0.2%) | 1,281 (0.2%) |  |  |
| Home ART delivery | 1,366 (0.4%) | 2,020 (0.3%) |  |  |
| Community ART distribution points | 2,693 (0.8%) | 4,989 (0.9%) |  |  |
| Adherence groups | 2,275 (0.7%) | 4,550 (0.8%) |  |  |
| *Health outcomes [11]* |  |  |  |  |
| Retention rates | Outcomes dependent on DSD model type, age, sex and setting (urban/rural). |  |  |  |
| Viral suppression rates |  |  |  |  |
| *Costs per year (mean) -2023 USD* | *Average provider costs per client per year [17]* | *Average costs to clients per year [18]* |  |  |
|  |  | *Lost wages (opportunity costs)* | *Transport costs* |  |
| | | | % incurring any transport costs | Average cost per client per year (if >\$0) |
| Conventional care not eligible for DSD | \$119.23 | \$6.94 | 51.5% | \$3.21 |
| Conventional care (3MMD) | \$109.05 | \$6.59 | 40.9% | \$2.92 |
| Six-month dispensing (6MMD) | \$99.72 | \$3.01 | 36.4% | \$2.92 |
| Fast-track refills | \$105.90 | \$5.86 | 42.5% | \$1.25 |
| Extended clinic hours | \$109.05 | \$2.35 | 46.7% | \$1.74 |
| Scholar/adolescent model | \$109.05 | \$10.90 | 26.8% | \$1.75 |
| Mobile ART distribution | \$125.56 | \$4.01 | 0.0% | \$0.00 |
| Health post | \$107.27 | \$5.47 | 51.4% | \$1.71 |
| Home ART delivery | \$132.51 | \$4.57 | 40.0% | \$3.00 |
| Community ART distribution points | \$106.19 | \$5.47 | 51.4% | \$1.71 |
| Adherence groups | \$104.33 | \$11.07 | 34.8% | \$3.04 |

### Cost inputs

Provider costs using ingredients-based approach and costs to clients were estimated using previously published (Table 3) [17,19]. Provider costs per ART client per year included costs of staff time (facility visits and DSD interactions), medication costs, and laboratory testing costs (Table S2) [17]. Costs to clients are based on self-reported resource utilization collected during the SENTINEL survey [20]. Clients in each ART delivery models were asked to report the number of interactions (visits) with the healthcare system and the time spent accessing care at each study site [20]. Cost to clients included opportunity costs for time spent seeking care, estimated as time spent accessing care multiplied by Zambia’s minimum daily wage ($1.99), and transport costs incurred for accessing ART services [18], taking into consideration the proportion of clients incurring transport costs in each ART delivery model (Table S3). Unit costs for the provider and costs to ART clients per year were updated to 2023 prices [17,18,21] and then converted from Zambian Kwacha (ZMW) to United States Dollars (USD) using the average 2023 exchange rate of 20.23 ZMW per = 1 USD [21].

### Scenarios and input assumptions

As noted above, the base case was defined as the current national distribution of ART clients by age, sex, setting and ART delivery model type, under the current status quo, as determined by the SmartCare database (Table S1, Table 3). The analytic scenarios include 125 unique possible combinations of DSD models and model offerings tailored by age, sex, and setting. While most DSD models allow enrolment of any eligible ART patient, some are limited to specific population subgroups. These include the scholar/adolescent model (for ART clients aged ≤24 years), extended clinic hours (for clients aged ≥25 years), and mobile ART delivery (available only in rural areas). We thus modelled two different sets of scenarios: 1) DSD models for the full population (excluding DSD models that are targeted to specific population subgroups); and 2) stratified scenarios which included age-specific DSD models for relevant subgroups and other DSD models for everyone else. For scenarios with two or more DSD models, we assumed equal distribution of the number of eligible ART clients allocated to each model.

Each scenario reflected a different allocation of eligible clients to the models included in the scenario. In most scenarios, 95% of ART clients were allocated to differentiated models (non-conventional care), with the rest remaining in conventional care. For example, in a fast track refills-only scenario, all eligible clients were enrolled in fast-track refills and the remainder were in conventional care. In a population specific scenario such as enrolling everyone eligible for DSD in a combination of scholar/adolescent model and extended clinic hours, eligible clients aged ≤24 years were enrolled in scholar/adolescent model and those aged ≥25 years enrolled in extended clinic hours. The details of mix of ART delivery models and client distribution by scenario are described in Table S4.

### Cost analysis

ADAPT estimates costs over a one-year period after ART delivery model enrolment from the perspectives of the provider (Ministry of Health) and of clients enrolled in each model. The relatively short (1-year) time horizon was chosen because it matched the period of the health outcomes, and we wanted to ensure realistic estimates that could be used for short-term budget cycles relevant to the Ministry of Health’s budget planning. For baseline and modelled scenarios, health system costs per model of ART delivery was estimated by multiplying the number of ART clients assigned to each model by average annual provider cost. Cost to clients were estimated by multiplying the number of clients assigned to each ART delivery model by the annual average costs incurred accessing ART services, including lost wages (opportunity costs) and transport costs.

### Cost-effectiveness analysis

To estimate incremental cost effectiveness ratios, we first ranked total health system costs of each scenario from lowest to highest total costs [22]. We then determined and eliminated scenarios that were strongly dominated. A scenario was considered strongly dominated if it resulted in higher total costs to the healthcare system and fewer people virally suppressed on treatment to the next best scenario [22]. An ICER for each scenario was then calculated by dividing the difference in total health system costs (incremental cost) by the difference in viral suppression rate (incremental effect) of one scenario compared to the next best scenario. ICERs per additional person suppressed on treatment were calculated by comparing scenarios to the to the next best scenario scenarios that was on the cost-effective frontier [22].

Next, we compared each scenario with the next best scenario, based on its ICER, and scenarios that were weakly dominated were identified and eliminated. A scenario was considered weakly dominated if it resulted in smaller effect (lower number of people virally suppressed) but had a higher ICER compared to the next highest ranked scenario [22,23]. These results were then used to establish the cost-effectiveness frontier, defined as graphical representation of interventions that increase the number of people virally suppressed on treatment compared to the current distribution of ART delivery model [24]. A full list of all scenarios analysed, with total effects, total costs, and ICERs, is provided in Table S5. Finally, for all interventions on the cost-effectiveness frontier, a sub-analysis was conducted to understand how health outcomes (viral suppression and retention rates) of each sub-group (by age, sex, and rural/urban setting) would be affected if the scenarios on the cost-effectiveness frontier were adopted at the national level, compared to base case distribution.

## Results

### Base case scenario

The base case cohort comprised a total of 917,749 ART clients, of whom roughly two thirds were female (63.7%) and two thirds live in an urban setting (69.3%) (Table 3). Most (72%) were aged between 25-49 years and another quarter were ≥50 years, leaving just 8% ≤24 years. In the base case, reflecting the status quo in 2022, less than a quarter of the cohort (20%) received conventional care, nearly 70% 6MMD, 9% fast track, and the remainder were enrolled in each of the other active models, which served fewer than 1% of clients each. The average provider cost ranged from $100 to $133 per client per year, while costs to clients ranged from $3.25 and $11.07 per year for lost wages. Among the 39.2% who reported incurring any transport costs, cost per client ranged from $1.71 to $3.21 per year (Table 4).

**Table 4.** Health outcomes and costs for the Zambia ART program at 12 months under the base case, 2022.

| Characteristics | National outcomes, n, % |
| --- | --- |
| **Analytic cohort** |  |
| Total (N) | 917,749 (100%) |
| Retained in care | 817,948 (89.1%) |
| Suppressed on treatment | 770,086 (94.1%) |
| **Costs (2023 USD)** |  |
| Health system costs | \$84,332,234 (n/a) |
| Costs to clients | \$4,145,400 (n/a) |
| **DSD eligibility/enrolment†** |  |
| Conventional care not eligible for DSD | 48,679 (5.3%) |
| Conventional care eligible for DSD but not enrolled | 133,031 (14.5%) |
| Clients enrolled in DSD models | 736,039 (80.2%) |
| † Clients enrolled in DSD models and those eligible but not enrolled can be allocated to any DSD model for the scenario analysis; clients not eligible for DSD must remain in conventional care. |  |

A total of 89.1% (n=817,948) of the cohort were retained in care at 12 months in the base case scenario (Table 5). Among those retained in care, 94.1% (n=770,086) were virally suppressed. Of the entire cohort, 80.2% (n=736,039) were enrolled in DSD models, 14.5% (n=133,031) were eligible for DSD but not enrolled, and 5.3% (n=48,679) were not eligible for DSD enrolment. The associated health system costs and costs to clients were $84,332,234 and $4,145,400 per year, respectively.

**Table 5.** Health outcomes, health system costs, and ICERs for the DSD scenarios on the cost-effectiveness frontier.

| Scenarios | Description | Total number of people retained (n, % change compared to base case) | Total number of people suppressed (n, % change compared to base case) | Total health system cost (USD, % change) | ICER per additional person suppressed (USD)† |
| --- | --- | --- | --- | --- | --- |
| Base case | Current distribution of ART delivery models as indicated in the Smartcare database (80.2% enrolled in DSD, 14.5% enrolled in conventional care and eligible for DSD but not enrolled, and 5.3% in conventional care but not eligible for DSD). | 817,948 (n/a) | 770,086 (n/a) | \$84,332,234 (n/a) | n/a |
| 6MMD | 94.7% (n=869,070) of all ART clients enrolled in 6MMD across all population groups (rural/urban, age, sex) (all of those eligible for DSD models), and 5.3% (n= 48,679) remains in conventional care (not eligible for DSD models). | 827,415 (1.2%) | 782,545 (1.6%) | \$83 095,136 (-1.5%) | Cost-saving (compared to base case) |
| 6MMD & AGs | 94.7% (n=869,070) of all ART clients distributed equally between a combination of two DSD models (6MMD & AGs) and 5.3% (n= 48,679) remain in conventional care (not eligible for DSD models). | 831,498 (1.7%) | 791,712 (2.8%) | \$85,336,549 (1.2%) | \$245 |
| AGs | 94.7% (n=869,070) of all ART clients enrolled in AGs across all population groups (rural/urban, age, sex) and 5.3% (n= 48,679) remains in conventional care not eligible. | 835,581 (2.2%) | 800,878 (4.0%) | \$87,577,961 (3.8%) | \$245 |
| FTRs & AGs | 94.7% (n=869,070) of all ART clients distributed equally between a combination of two DSD models (FTRs & AGs) across all population groups (rural/urban, age, sex) and 5.3% (n= 48,679) remain in conventional care (not eligible for DSD models). | 838,225 (2.5%) | 802,755 (4.2%) | \$88,483,273 (4.9%) | \$482 |
| FTRs | 94.7% (n=869,070) of all eligible clients enrolled in FTRs only across all population groups (rural/urban, age, sex) and 5.3% (n= 48,679) remains in conventional care (not eligible for DSD models). | 840,869 (2.8%) | 804,632 (4.5%) | \$89,388,584 (6.0%) | \$482 |
| AGs & HAD | Population specific-scenario for all 94.7% (n= 869,070) eligible clients; enrolling 6.8% (n= n=62,106) of clients aged ≤24 years in AGs and 87.7% (n= 806,964) of clients aged ≥25 years in HAD across all population groups (rural/urban, age, sex) and 5.3% (n= 48,679) of all clients remain in conventional care (not eligible for DSD models). | 844,609 (3.3%) | 807,356 (4.8%) | \$109,574,215 (29.9%) | \$7,409 |

† ICERs per additional person suppressed on treatment were calculated by comparing scenarios to the previous scenarios that was on the cost-effective frontier.
6MMD, six-month dispensing; AGs, adherence groups; FTRs, fast track refills; HAD, home ART delivery

### Scenarios on the cost-effectiveness frontier

Results of all 125 scenarios analysed are reported in Table S5. Six of these scenarios were found to be on the cost-effectiveness frontier (Table 5, Figure 2). Five of the scenarios on the frontier improved outcomes but also resulted in greater health system costs and costs to clients, while one scenario—6-month dispensing for all eligible clients—both improved outcomes and reduced costs, compared to the current baseline. Scenarios on the cost-effectiveness frontier encompassed both facility-based models (6MMD, fast-track refills) and out-of-facility-based models (adherence groups, home ART delivery) for all those eligible for DSD enrolment. The only scenario on the cost-effectiveness frontier that included an approach that targeted specific sub-populations was the combination of adherence groups (AGs) (for those ≤24 years) and home ART delivery (HAD) (for those aged ≥25).

**Figure 2.**
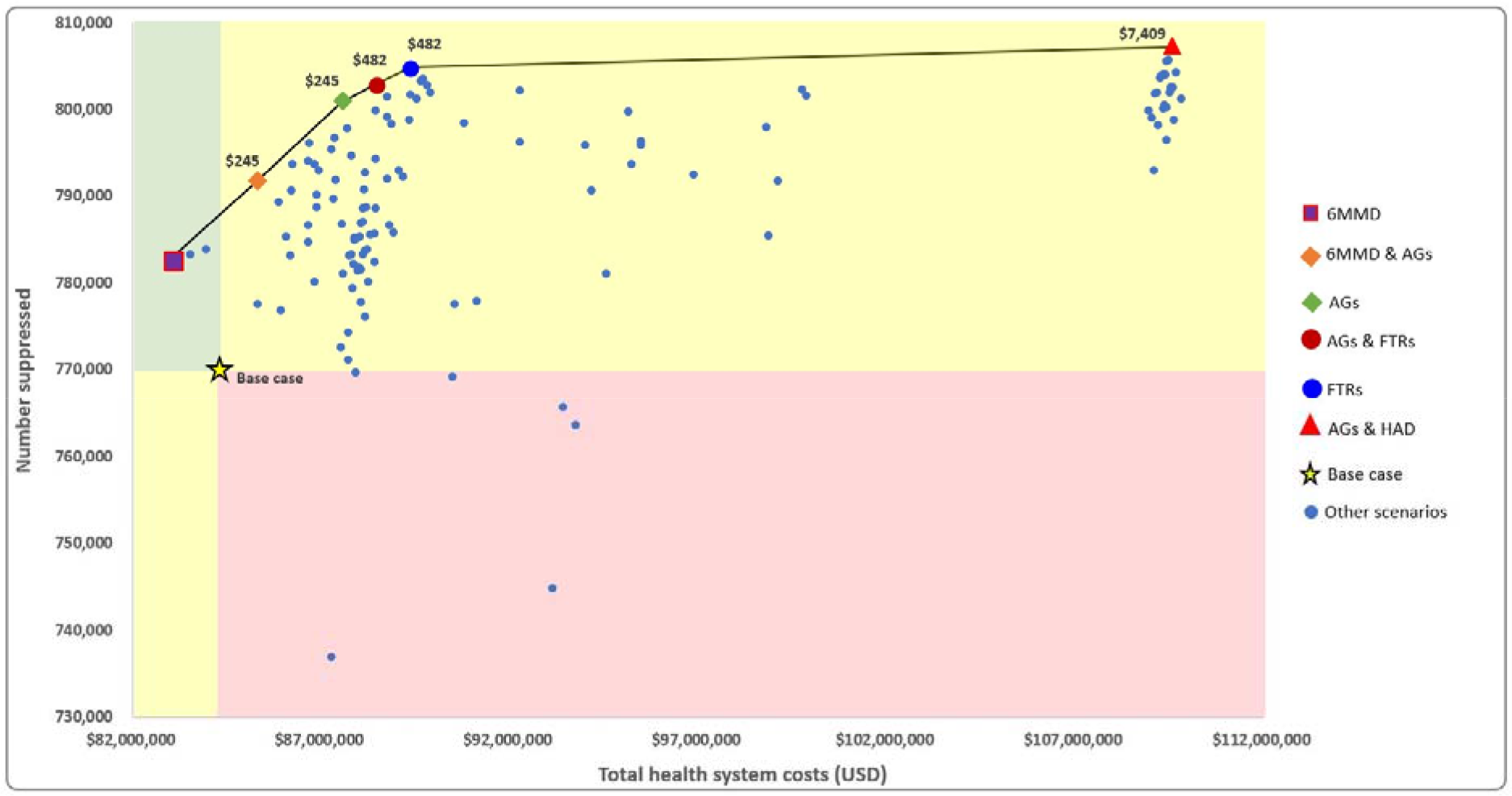
Total number of people suppressed on treatment by total health system costs for scenarios on the cost-effectiveness frontier

With the exception of 6-month dispensing, which was cost-saving, all other scenarios on the cost-effectiveness frontier resulted in an additional cost to the healthcare system per additional person virally suppressed, with ICERs ranging from $245 to $7,409. Compared to the base case, enrolling all eligible clients in 6MMD increased the number of individuals retained in care by 1.2% and the number virally suppressed by 1.6%, while decreasing health system cost and costs to clients by 1.5% and 16.6%, respectively. As illustrated in Figure 2, the remaining scenarios on the frontier further improved treatment outcomes while also increasing total costs. The most expensive scenario on the cost-effectiveness frontier—AGs for young adults and HAD for everyone else—resulted in the greatest increase in suppression (4.8% more than the base case), at an increase in annual costs of nearly 30% compared to the current base case.

### Costs to ART clients

Compared to the current base case ART distribution of DSD models, enrolling all clients eligible for DSD in 6MMD-only scenario was cost-saving, while the other five scenarios on the cost-effectiveness frontier all resulted in an increased cost to clients per year (Figure 3). Compared to the base case, enrolling all eligible clients in 6MMD-only reduced cost to clients by 16.6%, while a combination of 6MMD and AGs resulted in an increase in costs to clients per year of 63.0%. The scenario on the cost-effectiveness frontier with the highest cost to ART clients was AGs-only for all eligible clients, which increased costs to clients by 142.6% compared to the current base case. A combination of fast-track refills (FTRs) and AGs for all eligible clients resulted in an annual increase of costs to clients of 86.4% compared to the base case. Enrolling all eligible clients in FTRs-only, and the age-specific scenario, which was a combination of AGs for those ≤24 years and HAD for those ≥25 years, increased costs to clients per year by nearly 31% compared to the base case.

**Figure 3.**
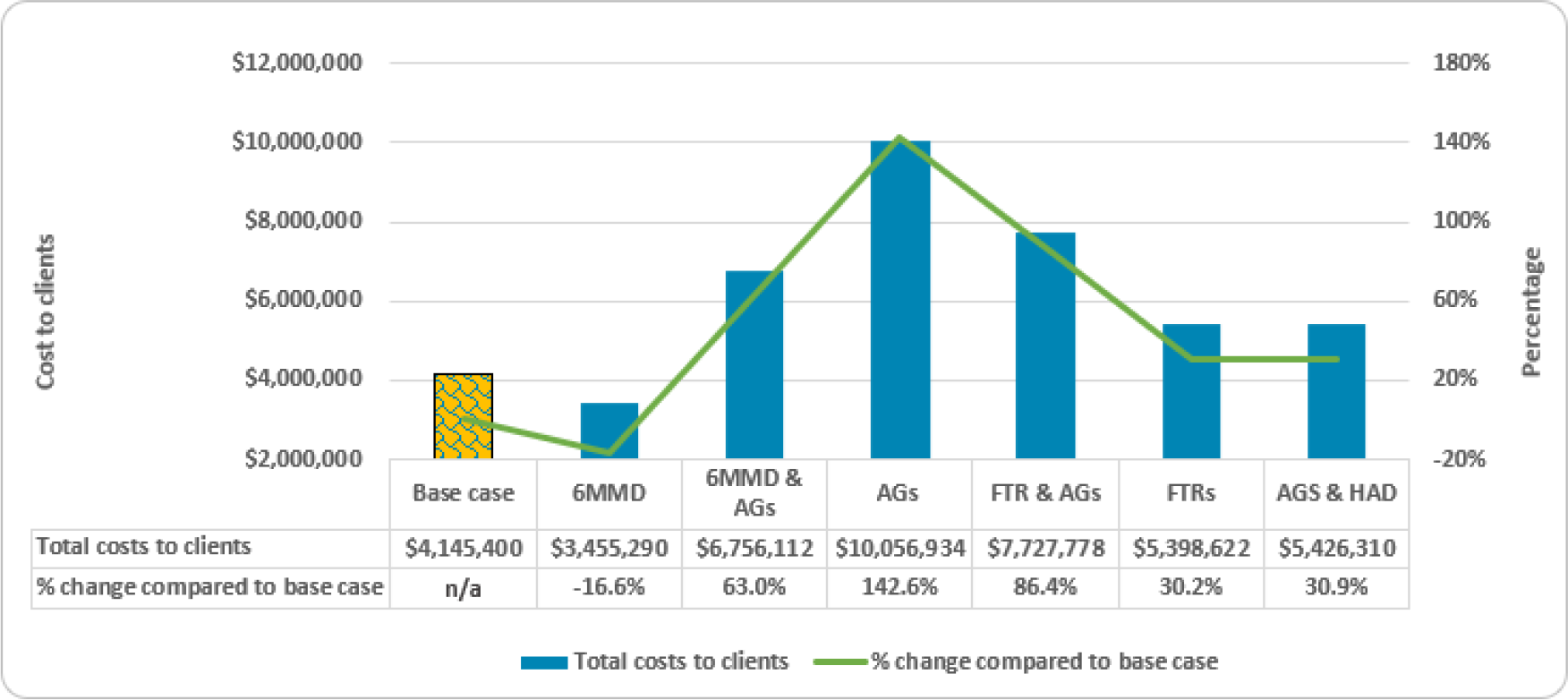
Cost to clients per year for the scenarios on the cost-effectiveness frontier (2023 USD)

### Outcomes stratified by subpopulation

The relative impact of each scenario varied by age, sex, and setting (Table 6). For clients aged 25-49 years, who constituted over 70% of the ART population included in the analysis, all scenarios on the cost-effectiveness frontier resulted in increased retention compared to the base case. Retention was lower in female clients aged 15-19 years and male clients 15-24 in rural settings for all scenarios on the cost-effectiveness frontier except for 6MMD-only, which improved retention across all sub-populations. A scenario utilizing AGs-only had a low retention rate among clients aged ≥50+ in both urban and rural settings. Viral suppression increased for most subpopulations in most scenarios on the cost-effectiveness frontier, though 6MMD was slightly less successful for older clients and rural clients showed slightly lower suppression rates in several of the scenarios. 6MMD combined with adherence groups was associated with better viral suppression for the largest proportion of the subpopulations assessed except for male clients aged ≥50+ in rural settings.

**Table 6.** Retention and viral suppression rates for scenarios on cost-effectiveness frontier compared to the base case distribution by ART client sub-category.

| a) |  | Retention rate (%) |  |  |  |  |  |  |
| --- | --- | --- | --- | --- | --- | --- | --- | --- |
| Sub-categories |  | Base case Distribution | 6MMD | 6MMD & AGs | AGs | FTRs & AGs | FTRs | AGs & HAD |
|  | Overall | 89.1% | 90.2% | 90.6% | 91.0% | 91.3% | 91.6% | 92.0% |
|  | Male, 15-19, Rural | 90.6% | 91.7% | 89.5% | 87.2% | 82.3% | 77.3% | 87.2% |
|  | Male, 20-24, Rural | 88.6% | 90.2% | 92.7% | 95.1% | 92.1% | 89.1% | 95.1% |
|  | Male, 25-49, Rural | 89.8% | 91.1% | 91.2% | 91.3% | 91.5% | 91.8% | 92.1% |
|  | Male, 50+, Rural | 91.7% | 92.5% | 91.2% | 89.9% | 92.5% | 95.2% | 90.3% |
|  | Male, 15-19, Urban | 87.8% | 88.8% | 87.8% | 86.8% | 87.1% | 87.4% | 86.8% |
|  | Male, 20-24, Urban | 85.7% | 87.5% | 91.0% | 94.5% | 91.5% | 88.5% | 94.5% |
|  | Male, 25-49, Urban | 87.8% | 88.9% | 90.0% | 91.1% | 91.2% | 91.4% | 89.9% |
|  | Male, 50+, Urban | 90.1% | 90.3% | 90.1% | 89.9% | 91.2% | 92.4% | 91.5% |
|  | Female, 15-19, Rural | 88.0% | 89.0% | 88.0% | 87.1% | 82.3% | 77.5% | 87.1% |
|  | Female, 20-24, Rural | 85.9% | 87.5% | 91.4% | 95.4% | 92.3% | 89.2% | 95.4% |
|  | Female, 25-49, Rural | 90.7% | 91.8% | 91.7% | 91.5% | 91.7% | 91.9% | 96.5% |
|  | Female, 50+, Rural | 92.4% | 93.1% | 91.5% | 90.0% | 92.6% | 95.3% | 85.7% |
|  | Female, 15-19, Urban | 86.3% | 87.7% | 87.1% | 86.5% | 86.8% | 87.1% | 86.5% |
|  | Female, 20-24, Urban | 82.6% | 85.5% | 89.5% | 93.4% | 90.5% | 87.6% | 93.4% |
|  | Female, 25-49, Urban | 88.8% | 89.8% | 90.5% | 91.2% | 91.4% | 91.6% | 93.9% |
|  | Female, 50+, Urban | 90.8% | 91.0% | 90.5% | 90.0% | 91.3% | 92.5% | 84.2% |

| b) |  | Suppression rate (%) |  |  |  |  |  |  |
| --- | --- | --- | --- | --- | --- | --- | --- | --- |
| Sub-categories |  | Base case Distribution | 6MMD | 6MMD & AGs | AGs | FTRs & AGs | FTRs | AGs & HAD |
|  | Overall | 94.1% | 94.6% | 95.2% | 95.8% | 95.8% | 95.7% | 95.6% |
|  | Male, 15-19, Rural | 80.3% | 81.9% | 86.7% | 91.8% | 91.3% | 90.7% | 91.8% |
|  | Male, 20-24, Rural | 84.6% | 87.3% | 89.6% | 91.7% | 91.3% | 90.9% | 91.7% |
|  | Male, 25-49, Rural | 94.3% | 95.1% | 94.6% | 94.0% | 93.7% | 93.3% | 92.6% |
|  | Male, 50+, Rural | 95.3% | 95.7% | 95.0% | 94.2% | 93.8% | 93.4% | 92.8% |
|  | Male, 15-19, Urban | 78.5% | 78.9% | 85.8% | 92.8% | 92.9% | 92.9% | 92.8% |
|  | Male, 20-24, Urban | 80.5% | 80.6% | 87.2% | 93.4% | 93.3% | 93.3% | 93.4% |
|  | Male, 25-49, Urban | 93.9% | 94.2% | 95.0% | 95.8% | 95.8% | 95.9% | 96.9% |
|  | Male, 50+, Urban | 94.8% | 94.7% | 95.4% | 96.0% | 96.1% | 96.1% | 97.2% |
|  | Female, 15-19, Rural | 80.2% | 82.0% | 87.8% | 93.7% | 93.1% | 92.4% | 93.7% |
|  | Female, 20-24, Rural | 91.5% | 93.4% | 94.6% | 95.8% | 95.3% | 94.7% | 95.8% |
|  | Female, 25-49, Rural | 95.2% | 95.8% | 96.2% | 96.6% | 96.0% | 95.5% | 92.7% |
|  | Female, 50+, Rural | 96.3% | 96.5% | 96.6% | 96.8% | 96.2% | 95.7% | 92.8% |
|  | Female, 15-19, Urban | 78.4% | 79.3% | 86.0% | 92.7% | 92.9% | 93.0% | 92.7% |
|  | Female, 20-24, Urban | 90.0% | 91.8% | 93.4% | 94.9% | 95.0% | 95.1% | 94.9% |
|  | Female, 25-49, Urban | 94.9% | 95.2% | 95.7% | 96.2% | 96.4% | 96.5% | 97.0% |
|  | Female, 50+, Urban | 95.7% | 95.5% | 96.0% | 96.5% | 96.6% | 96.7% | 97.3% |

■ > Base case ■ < Base case

## Discussion

In this modelling study, we identified several differentiated service delivery allocation scenarios that are likely to have better health outcomes than the current distribution of ART delivery models in Zambia and estimated the additional cost to the HIV program budget of implementing those scenarios. Enrolment of all eligible clients in 6MMD appeared to be cost-saving for both providers and clients, while still improving outcomes modestly. While not cost-saving, other combinations of DSD models on the cost-effectiveness frontier could further increase retention in care and viral suppression, beyond rates estimated for 6MMD-only, by up to 4.8%. These scenarios come at an additional cost to both providers and clients, compared to the base case. This is not surprising in view of the base case distribution, in which nearly 70% of clients were enrolled in 6MMD. Since all other models of care are more expensive for both providers and clients than is 6MMD, any scenario that reduced total 6MMD coverage was bound to cost more than the base case. By increasing viral suppression, however, the combinations of DSD models on the frontier may have the longer-term benefit of reducing onward transmission, and therefore reducing both health system and client costs in the long term.

Our analysis found that a scenario tailored to specific sub-populations–adherence groups for those ≤24 years and home ART delivery for those ≥25+ years–can increase the number of people virally suppressed on treatment across all subgroups. In the 6MMD-only scenario, in contrast, retention and suppression rates increased for all subgroups except clients ≥50+ years in urban settings. DSD models tailored to subpopulations may thus still be required, rather than implementation of a single DSD scale-up strategy for all clients who are eligible.

To our knowledge, this is the first model to consider how the distribution of available differentiated service delivery models might affect overall cost-effectiveness of a national HIV treatment program. Other studies have investigated the outcomes, costs, and cost-effectiveness of individual DSD models, with a wide range of results depending on the country and specific model(s) of care being evaluated [7,25,26], or have compared the costs of the actual DSD landscape to a “no DSD” status quo[10]. Across multiple countries, reported retention and viral suppression rates in DSD models have broadly fallen within 5% of what was reported in the conventional care [2]. A retrospective electronic record review in Zambia reported higher cost of community DSD models compared to conventional care, with an annual provider cost per DSD ranged from $116 to $199 for the DSD models, compared to $100 in conventional care [17]. A pragmatic, cluster-randomised, unblinded, non-inferiority trial conducted in Zambia and Malawi showed that 6-month ART dispensing was associated with non-inferior retention rates and ART uptake to standard of care and 3-month ART dispensing [27]. In this trial, as in our modelled analysis, 6-month ART dispensing was found to be cost-saving for both healthcare system and to clients retained in care at 12 months, compared to standard of care and 3-month ART dispensing [27]. In two prospective cluster randomized non-inferiority trials conducted in Lesotho and Zimbabwe, community adherence groups with 3 months medication dispensing and 6-month community dispensing were found to be less costly than conventional care [28,29], with similar health outcomes. Finally, a comprehensive analysis of the costs of existing DSD models in Mozambique concluded that, compared to a hypothetical scenario in which all clients received conventional care, DSD is less expensive than the comparison[10]. These studies support our finding that DSD models have a potential to improve health outcomes and minimise costs.

Although our results are specific to one country and point in time, we believe that they can broadly help inform policy-makers about how changes in DSD model offerings might affect health outcomes, costs, and the prioritisation of population specific DSD model combinations at an aggregate level and the trade-offs that must be made. If the goal of the healthcare system is to achieve optimal cost savings without diminishing health outcomes, for example, the implementation of 6MMD-only for all eligible ART clients appears to be a strategy that could achieve such goals. If the goal is to prioritise improvement of health outcomes while allowing the budget to increase modestly, scenarios that increase health outcomes but with lower ICERs may be optimal. The higher costs to clients associated with most non-conventional models of care, along with client preferences and concerns, must also be considered. Future research is required to understand the DSD model transition acceptability and cost implications from the perspective of the clients.

Key strengths of our study are the use of a large national-level dataset to quantify the expected outcomes of different DSD models in different sub-populations, local cost estimates, and the analysis of multiple hypothetical combinations of DSD models. Our study also had a number of limitations. First, we parameterised the model using retention and viral suppression rates from individuals enrolled in these models currently. Our results omit the effects and costs of moving from the current distribution to any other scenario. Moving individuals around among models, as is done in our analysis, is likely to result in different outcomes and costs from those observed for current enrollees, particularly if the base case reflects individual client preferences for service delivery, which the modelled scenarios ignore. For this reason, results should be regarded as illustrative of what can potentially be achieved through re-allocation, rather than a precise prediction of outcomes.

Second, we modelled only national level outcomes with stratification for rural and urban settings. We did not consider protentional provincial or other geographic differences and we acknowledge that the results may differ at provincial level especially between provincial with substantial difference in geographical settings and poor access to care. Third, because we did not have actual cost estimates for all the models of care included in our analysis, we assumed that models are implemented according to guidelines and utilized ingredients-based costing. Actual costs incurred may be either under-or over-estimated using this approach. Fourth, cost to clients were estimated from self-reported frequency of visits, time spent at the facility, lost wages, and transport costs. Self-report may be subject to recall bias, reducing the accuracy of estimates. Fifth, we did not include any additional costs that might arise from the additional coordination and management required to operate multiple models of care at a single site, nor did we capture any costs above those at the facility (e.g. national or provincial planning, training, budgeting). Some facilities are likely to struggle with the simultaneous implementation of multiple models and require additional resources to do this successfully. We note that none of the scenarios on the cost-effectiveness frontier, however, required more than two non-conventional models of care, reducing our concerns about imposing excessive management and coordination burden.

Finally, we acknowledge that the results of this modelling analysis may not be generalizable beyond Zambia, as the factors that influence the cost-effectiveness of DSD allocation may be context dependent. For countries in southern Africa with relatively similar conditions as Zambia, the results we report may be adoptable locally. For most countries, though, it is the mathematical model itself, and the potential for running it with country-specific data, rather than using the Zambia-specific results, that we hope can help improve DSD allocation. We also emphasize that “cost savings” in a model like that presented here in fact reflect a reduced investment of resources, such as staff time, per ART client served, rather than a budgetary savings. The freed-up resources can be used for other purposes within the healthcare system, but budgetary reductions should not be expected.

## Conclusions

With six years remaining towards achieving the global UNAIDS 95-95-95 targets in 2030, many countries in sub-Saharan Africa, such as Zambia, that have made progress towards achieving these goals may benefit from restructuring their DSD model offerings for ART distribution [30]. The model and overall approach demonstrated here can broadly allow for rapid evaluation of different model allocations in terms of health outcomes, cost to clients, and costs to the healthcare system, while ensuring that all client sub-populations are considered. Given the large scale of ART programs and the widespread adoption of differentiated service delivery in sub-Saharan Africa, even small improvements in service delivery efficiency can have large impacts. Applying a relatively simple mathematical model that relies largely on existing data may be one way to achieve such progress.

## Ethics

This study protocol was approved by ERES Converge IRB (Zambia), protocol number 2019-Sep-030; the Human Research Ethics Committee (Medical) of the University of Witwatersrand (South Africa), protocol number M190453; and the Boston University IRB (United States), protocol number H-38823.

## Authors’ contributions

NL, SR, LJ, BP and BEN conceptualised the study. NL and LJ collected the cost data. LJ and BEN oversaw data collection. NL, LJ and BEN developed and parameterised the model. NL, LJ and BEN did data analysis and had full access to the data. NL, SR, LJ and BEN verified underlying data, assumptions and drafted manuscript. All authors contributed to reviewing and editing the manuscript for submission.

## Data Availability

Non-human subjects data are contained in the manuscript. Human subjects data are owned by the Zambian Ministry of Health and their use was approved by the ERES Converge IRB (Zambia). Full data are available upon approval from the Zambian Ministry of Health and appropriate ethics committees.

## Acknowledgements

We would like to thank the staff of the Ministry of Health in Zambia for approving this research. We are particularly appreciative of the support of each country’s Differentiated Service Delivery Technical Working Group.

## Supplementary files

Table S1. Health outcomes – retention and viral suppression rates stratified by ART delivery model, sex, setting and age group

Table S2. Unit costs (provider costs)

Table S3. Average cost (2023 USD) to ART clients per year, stratified by ART delivery model

Table S4. Distribution of clients among ART delivery models for the scenarios analysed

Table S5. Cost-effectiveness analysis of all scenarios

## List of abbreviations

DSD: Differentiated service delivery
ART: Antiretroviral therapy
ICER: Incremental cost-effectiveness ratio
6MMD: Six-months dispensing
FTRs: Fast track refills
AGs: Adherence groups
HAD: Home ART delivery

