## Supplementary files for "Optimal mix of differentiated service delivery models for HIV treatment in Zambia: a mathematical modelling study"

### Table S1. Health outcomes – retention and viral suppression rates stratified by ART delivery model, sex, setting and age group

The distribution of ART delivery model which reflect the 2022 cohort by sex, settings, age and health outcomes (retention rates and suppression rates) were estimated from the Zambia electronic medical record – Smartcare database [1].

| **ART delivery model** | **Sex** | **Setting** | **Age** | **Retention rates** | **Suppression rates** |
| --- | --- | --- | --- | --- | --- |
| Scholar/adolescents model | Female | Rural | 15-19 | 94.4% | 91.7% |
| Fast track refills | Female | Rural | 15-19 | 76.5% | 95.7% |
| Conventional care not eligible for DSD | Female | Rural | 15-19 | 81.9% | 67.6% |
| Six-month dispensing (6MMD) | Female | Rural | 15-19 | 89.6% | 83.3% |
| Adherence groups | Female | Rural | 15-19 | 87.5% | 96.8% |
| Community ART distribution points | Female | Rural | 15-19 | 95.0% | 75.0% |
| Health posts | Female | Rural | 15-19 | 91.5% | 100.0% |
| Mobile ART distribution | Female | Rural | 15-19 | 94.9% | 92.0% |
| Conventional care (3MMD) | Female | Rural | 15-19 | 87.4% | 76.2% |
| Scholar/adolescents model | Male | Rural | 15-19 | 90.0% | 91.7% |
| Fast track refills | Male | Rural | 15-19 | 76.5% | 93.5% |
| Conventional care not eligible for DSD | Male | Rural | 15-19 | 80.7% | 63.9% |
| Six-month dispensing (6MMD) | Male | Rural | 15-19 | 92.5% | 83.2% |
| Adherence groups | Male | Rural | 15-19 | 87.5% | 94.2% |
| Community ART distribution points | Male | Rural | 15-19 | 95.0% | 75.0% |
| Health posts | Male | Rural | 15-19 | 89.9% | 92.9% |
| Mobile ART distribution | Male | Rural | 15-19 | 95.7% | 87.5% |
| Conventional care (3MMD) | Male | Rural | 15-19 | 90.4% | 73.1% |
| Scholar/adolescents model | Female | Urban | 15-19 | 93.0% | 83.3% |
| Fast track refills | Female | Urban | 15-19 | 88.2% | 96.8% |
| Conventional care not eligible for DSD | Female | Urban | 15-19 | 77.3% | 66.0% |
| Six-month dispensing (6MMD) | Female | Urban | 15-19 | 88.9% | 80.9% |
| Adherence groups | Female | Urban | 15-19 | 87.5% | 96.5% |
| Community ART distribution points | Female | Urban | 15-19 | 95.0% | 75.0% |
| Health posts | Female | Urban | 15-19 | 89.0% | 93.7% |
| Conventional care (3MMD) | Female | Urban | 15-19 | 86.0% | 74.4% |
| Scholar/adolescents model | Male | Urban | 15-19 | 96.7% | 83.3% |
| Fast track refills | Male | Urban | 15-19 | 88.2% | 96.2% |
| Conventional care not eligible for DSD | Male | Urban | 15-19 | 79.2% | 63.3% |
| Six-month dispensing (6MMD) | Male | Urban | 15-19 | 89.7% | 80.4% |
| Adherence groups | Male | Urban | 15-19 | 87.5% | 96.1% |
| Community ART distribution points | Male | Urban | 15-19 | 95.0% | 75.0% |
| Health posts | Male | Urban | 15-19 | 87.0% | 95.7% |
| Conventional care (3MMD) | Male | Urban | 15-19 | 87.8% | 74.7% |
| Scholar/adolescents model | Female | Rural | 20-24 | 94.4% | 91.7% |
| Fast track refills | Female | Rural | 20-24 | 90.4% | 95.7% |
| Conventional care not eligible for DSD | Female | Rural | 20-24 | 74.6% | 82.7% |
| Six-month dispensing (6MMD) | Female | Rural | 20-24 | 88.5% | 94.2% |
| Adherence groups | Female | Rural | 20-24 | 97.2% | 96.8% |
| Community ART distribution points | Female | Rural | 20-24 | 88.9% | 83.3% |
| Health posts | Female | Rural | 20-24 | 91.5% | 100.0% |
| Mobile ART distribution | Female | Rural | 20-24 | 94.9% | 92.0% |
| Conventional care (3MMD) | Female | Rural | 20-24 | 84.0% | 87.1% |
| Scholar/adolescents model | Male | Rural | 20-24 | 90.0% | 91.7% |
| Fast track refills | Male | Rural | 20-24 | 90.4% | 93.5% |
| Conventional care not eligible for DSD | Male | Rural | 20-24 | 75.9% | 63.0% |
| Six-month dispensing (6MMD) | Male | Rural | 20-24 | 91.6% | 89.4% |
| Adherence groups | Male | Rural | 20-24 | 97.2% | 94.2% |
| Community ART distribution points | Male | Rural | 20-24 | 88.9% | 83.3% |
| Health posts | Male | Rural | 20-24 | 89.9% | 92.9% |
| Mobile ART distribution | Male | Rural | 20-24 | 95.7% | 87.5% |
| Conventional care (3MMD) | Male | Rural | 20-24 | 84.7% | 76.4% |
| Scholar/adolescents model | Female | Urban | 20-24 | 93.0% | 83.3% |
| Fast track refills | Female | Urban | 20-24 | 90.3% | 96.8% |
| Conventional care not eligible for DSD | Female | Urban | 20-24 | 64.9% | 78.7% |
| Six-month dispensing (6MMD) | Female | Urban | 20-24 | 87.9% | 93.1% |
| Adherence groups | Female | Urban | 20-24 | 97.2% | 96.5% |
| Community ART distribution points | Female | Urban | 20-24 | 88.9% | 83.3% |
| Health posts | Female | Urban | 20-24 | 89.0% | 93.7% |
| Mobile ART distribution | Female | Urban | 20-24 | 94.3% | 96.5% |
| Conventional care (3MMD) | Female | Urban | 20-24 | 80.0% | 86.1% |
| Scholar/adolescents model | Male | Urban | 20-24 | 96.7% | 83.3% |
| Fast track refills | Male | Urban | 20-24 | 90.3% | 96.2% |
| Conventional care not eligible for DSD | Male | Urban | 20-24 | 72.6% | 69.3% |
| Six-month dispensing (6MMD) | Male | Urban | 20-24 | 89.1% | 81.8% |
| Adherence groups | Male | Urban | 20-24 | 97.2% | 96.1% |
| Community ART distribution points | Male | Urban | 20-24 | 88.9% | 83.3% |
| Health posts | Male | Urban | 20-24 | 87.0% | 95.7% |
| Conventional care (3MMD) | Male | Urban | 20-24 | 81.8% | 74.1% |
| Extended clinic hours | Female | Rural | 25-49 | 82.4% | 96.7% |
| Fast track refills | Female | Rural | 25-49 | 92.3% | 95.7% |
| Conventional care not eligible for DSD | Female | Rural | 25-49 | 80.5% | 90.1% |
| Six-month dispensing (6MMD) | Female | Rural | 25-49 | 92.2% | 96.0% |
| Adherence groups | Female | Rural | 25-49 | 91.9% | 96.8% |
| Community ART distribution points | Female | Rural | 25-49 | 92.1% | 95.9% |
| Health posts | Female | Rural | 25-49 | 91.5% | 100.0% |
| Home ART delivery | Female | Rural | 25-49 | 97.1% | 92.8% |
| Mobile ART distribution | Female | Rural | 25-49 | 94.9% | 92.0% |
| Conventional care (3MMD) | Female | Rural | 25-49 | 87.5% | 92.4% |
| Extended clinic hours | Male | Rural | 25-49 | 98.1% | 100.0% |
| Fast track refills | Male | Rural | 25-49 | 92.3% | 93.5% |
| Conventional care not eligible for DSD | Male | Rural | 25-49 | 77.7% | 88.6% |
| Six-month dispensing (6MMD) | Male | Rural | 25-49 | 91.7% | 95.3% |
| Adherence groups | Male | Rural | 25-49 | 91.9% | 94.2% |
| Community ART distribution points | Male | Rural | 25-49 | 92.1% | 95.9% |
| Health posts | Male | Rural | 25-49 | 89.9% | 92.9% |
| Home ART delivery | Male | Rural | 25-49 | 92.6% | 92.8% |
| Mobile ART distribution | Male | Rural | 25-49 | 95.7% | 87.5% |
| Conventional care (3MMD) | Male | Rural | 25-49 | 86.0% | 91.6% |
| Extended clinic hours | Female | Urban | 25-49 | 82.4% | 96.7% |
| Fast track refills | Female | Urban | 25-49 | 92.2% | 96.8% |
| Conventional care not eligible for DSD | Female | Urban | 25-49 | 76.1% | 89.7% |
| Six-month dispensing (6MMD) | Female | Urban | 25-49 | 90.4% | 95.4% |
| Adherence groups | Female | Urban | 25-49 | 91.9% | 96.5% |
| Community ART distribution points | Female | Urban | 25-49 | 92.1% | 95.9% |
| Health posts | Female | Urban | 25-49 | 89.0% | 93.7% |
| Home ART delivery | Female | Urban | 25-49 | 94.7% | 97.3% |
| Conventional care (3MMD) | Female | Urban | 25-49 | 86.7% | 93.4% |
| Extended clinic hours | Male | Urban | 25-49 | 98.1% | 100.0% |
| Fast track refills | Male | Urban | 25-49 | 92.2% | 96.2% |
| Conventional care not eligible for DSD | Male | Urban | 25-49 | 74.8% | 88.1% |
| Six-month dispensing (6MMD) | Male | Urban | 25-49 | 89.5% | 94.5% |
| Adherence groups | Male | Urban | 25-49 | 91.9% | 96.1% |
| Community ART distribution points | Male | Urban | 25-49 | 92.1% | 95.9% |
| Health posts | Male | Urban | 25-49 | 87.0% | 95.7% |
| Home ART delivery | Male | Urban | 25-49 | 90.7% | 97.3% |
| Conventional care (3MMD) | Male | Urban | 25-49 | 84.5% | 92.0% |
| Extended clinic hours | Female | Rural | 50+ | 82.4% | 96.7% |
| Fast track refills | Female | Rural | 50+ | 95.5% | 95.7% |
| Conventional care not eligible for DSD | Female | Rural | 50+ | 81.6% | 93.2% |
| Six-month dispensing (6MMD) | Female | Rural | 50+ | 93.2% | 96.5% |
| Adherence groups | Female | Rural | 50+ | 90.1% | 96.8% |
| Community ART distribution points | Female | Rural | 50+ | 91.8% | 99.5% |
| Health posts | Female | Rural | 50+ | 91.5% | 100.0% |
| Home ART delivery | Female | Rural | 50+ | 85.7% | 92.8% |
| Mobile ART distribution | Female | Rural | 50+ | 94.9% | 92.0% |
| Conventional care (3MMD) | Female | Rural | 50+ | 88.9% | 94.4% |
| Extended clinic hours | Male | Rural | 50+ | 98.1% | 100.0% |
| Fast track refills | Male | Rural | 50+ | 95.5% | 93.5% |
| Conventional care not eligible for DSD | Male | Rural | 50+ | 80.1% | 91.2% |
| Six-month dispensing (6MMD) | Male | Rural | 50+ | 92.7% | 95.8% |
| Adherence groups | Male | Rural | 50+ | 90.1% | 94.2% |
| Community ART distribution points | Male | Rural | 50+ | 91.8% | 99.5% |
| Health posts | Male | Rural | 50+ | 89.9% | 92.9% |
| Home ART delivery | Male | Rural | 50+ | 90.5% | 92.8% |
| Mobile ART distribution | Male | Rural | 50+ | 95.7% | 87.5% |
| Conventional care (3MMD) | Male | Rural | 50+ | 88.2% | 93.3% |
| Extended clinic hours | Female | Urban | 50+ | 82.4% | 96.7% |
| Fast track refills | Female | Urban | 50+ | 92.6% | 96.8% |
| Conventional care not eligible for DSD | Female | Urban | 50+ | 83.5% | 93.8% |
| Six-month dispensing (6MMD) | Female | Urban | 50+ | 91.1% | 95.5% |
| Adherence groups | Female | Urban | 50+ | 90.1% | 96.5% |
| Community ART distribution points | Female | Urban | 50+ | 91.8% | 99.5% |
| Health posts | Female | Urban | 50+ | 89.0% | 93.7% |
| Home ART delivery | Female | Urban | 50+ | 84.2% | 97.3% |
| Conventional care (3MMD) | Female | Urban | 50+ | 89.6% | 95.0% |
| Extended clinic hours | Male | Urban | 50+ | 98.1% | 100.0% |
| Fast track refills | Male | Urban | 50+ | 92.6% | 96.2% |
| Conventional care not eligible for DSD | Male | Urban | 50+ | 81.8% | 91.7% |
| Six-month dispensing (6MMD) | Male | Urban | 50+ | 90.4% | 94.8% |
| Adherence groups | Male | Urban | 50+ | 90.1% | 96.1% |
| Community ART distribution points | Male | Urban | 50+ | 91.8% | 99.5% |
| Health posts | Male | Urban | 50+ | 87.0% | 95.7% |
| Home ART delivery | Male | Urban | 50+ | 91.7% | 97.3% |
| Conventional care (3MMD) | Male | Urban | 50+ | 88.5% | 93.5% |

Table S2. Unit costs (provider costs)

| **Cost Items** | **Unit cost**  **(2023 USD)** | **Description** |
| --- | --- | --- |
| **Laboratory (cost per test)** |  | Assumed one test per person per year [2] |
| Viral load | $27.44 |  |
| CD4 count | $5.38 |  |
| **HIV treatment** |  | Assumed first-line regimen (Tenofovir/Lamivudine/  Dolutegravir) for all clients [2] |
| ART (per month) | $5.25 |  |
| **Facility visit (cost per visit)** |  | Unit cost data were collected from previously published literature in Zambia and updated with 2023 public sector prices and staff costs [3]. Facility visit and DSD interaction costs include staff time, equipment, consumables, overheads, and training costs [3]. |
| Clinical follow-up | $2.71 |  |
| Short clinic visit | $1.93 |  |
| Pharmacy visit | $0.59 |  |
| **DSD interaction (cost per interaction)** |  |  |
| Scholar/adolescent model | $1.11 |  |
| Fast-track refills | $1.93 |  |
| Adherence groups^†^ | $1.73 |  |
| Community ART distribution points‡ | $2.35 |  |
| Health post§ | $2.71 |  |
| Home ART delivery | $11.13 |  |
| Mobile ART distribution | $8.81 |  |
| † assumed similar resources use as Community Adherence groups and urban adherence groups  ‡ assumed similar resources use as urban adherence groups  § assumed similar resources use as clinical follow-up | | |

Table S3. Average cost (2023 USD) to ART clients per year, stratified by ART delivery model

| **Model of care** | **N** | **Number of health system interactions/RoC/year**  **(mean, SD)** | | | | | | **Opportunity cost/RoC/year† (mean, SD)** | | **Transport costs/RoC/year (mean, SD)** | |
| --- | --- | --- | --- | --- | --- | --- | --- | --- | --- | --- | --- |
|  |  | **Facility visits*** | | **Out-of Facility events** | | **Total** | | **Time spent (hours)** | **Cost** | **% RoC incurring any transport costs**** | **Travel costs/RoC incurring any transport costs** |
| Conventional care not eligible for DSD | 66 | 5.4 | (3.7) | - | - | 5.4 | (3.7) | 27.9 | $6.94 | 51.5% | $3.21 |
| Conventional care (3MMD) | 66 | 4.2 | (1.0) | 0.1 | (0.7) | 4.3 | (1.3) | 20.28 | $6.59 | 40.9% | $2.92 |
| Six-month dispensing | 118 | 2.2 | (0.8) | 0.2 | (1.1) | 2.4 | (1.4) | 12.16 | $3.01 | 36.4% | $2.80 |
| Scholar/adolescent model | 41 | 3.7 | (2.1) | 0.6 | (2.2) | 4.3 | (3.0) | 17.55 | $10.90 | 26.8% | $1.75 |
| Adherence groups | 23 | 2.0 | (1.0) | 4.6 | (4.1) | 6.7 | (4.2) | 26.06 | $11.07 | 34.8% | $3.04 |
| Community ART distribution points‡ | 37 | 2.6 | (1.4) | 3.4 | (3.8) | 6.0 | (3.5) | 21.10 | $5.47 | 51.4% | $1.71 |
| Extended clinic hours | 15 | 2.7 | (1.2) | 0.0 | (0.0) | 2.7 | (1.2) | 9.52 | $2.35 | 46.7% | $1.74 |
| Fast-track refills | 31 | 3.2 | (1.5) | 1.0 | (2.8) | 4.2 | (3.3) | 15.96 | $5.86 | 45.2% | $1.25 |
| Mobile ART distribution | 9 | 1.7 | (2.0) | 3.3 | (3.7) | 5.0 | (2.6) | 16.23 | $4.01 | 0.0% | $0.00 |
| Home ART delivery | 25 | 2.1 | (1.1) | 0.6 | (1.1) | 2.8 | (1.4) | 10.82 | $4.57 | 40.0% | $3.00 |
| Health post | 37 | 2.6 | (1.4) | 3.4 | (3.8) | 6.0 | (3.5) | 21.10 | $5.47 | 51.4% | $1.71 |
| Cost to clients data was collected and updated from previously collected data [4].  *Includes both clinical consult visits and ART medication pick up visits at the facility.  **Remainder likely walked, incurred no cash costs.  †Calculated opportunity cost based on the cost of time spent travelling and time at clinic visits or out-of-facility events. The minimum wage for Zambia of $1.98/day (adjusted to an hourly minimum wage using 8 working hours/day) as used to assign a monetary value to the time spent.  ‡Community ART distribution points are equivalent to external medication pickup points. | | | | | | | | | | | |

### Table S4. Distribution of clients among ART delivery models for the scenarios analysed

| Scenario number | Scenario name | Descriptions/DSD mix and distribution of ART clients |
| --- | --- | --- |
| Base case | 2022 ART distribution | The current ART program in Zambia which includes 10 ART delivery models plus conventional care (3MMD and frequent refills) distribution as indicated in 2022 SmartCare database.   - 80.2% DSD coverage, 14.1% in conventional care eligible for DSD but not enrolled, and 5.3% in conventional care not eligible for DSD. |
| *Scenario 1-4: The scenarios tailored to clients based on model population-specific enrolment with different DSD coverage (e.g. scholar/adolescent model is tailored for clients aged ≤24 years).* | | |
| 1 | Scholar/adolescent model-only | Enrolling all eligible clients aged ≤24 years in scholar/adolescent model   - 6.8% DSD coverage for clients aged ≤24 years, 90.1% conventional care (3MMD), and 3.1% remain in conventional care not eligible |
| 2 | MAD-only | Enrolling all eligible clients in rural settings in mobile ART delivery   - 29.3% DSD coverage of all clients in rural settings, 70.7% in conventional care (3MMD), and 3.1% remain in conventional care not eligible |
| 3 | HAD-only | Enrolling all eligible clients aged ≥25 years in Home ART delivery   - 87.9% DSD coverage for clients aged ≥25+, 9.0% conventional care (3MMD), and 3.1% remains in conventional care not eligible for DSD |
| 4 | ECH-only | Enrolling all eligible clients aged ≥25 years in extended clinic hours   - 87.9% DSD coverage for clients aged ≥25+, 9.0% conventional care (3MMD), and 3.1% remains in conventional care not eligible for DSD |
| *Scenario 5-9: Scenarios for all eligible clients, utilising DSD models with no age/setting specific restrictions, enrolling all clients in one DSD model at a time. Clients distribution: 94.7% DSD coverage and 5.3% conventional care not eligible for DSD.* | | |
| 5 | FTRs-only | Enrolling all eligible clients in fast-track refills |
| 6 | 6MMD-only | Enrolling all eligible clients in 6MMD |
| 7 | CADP-only | Enrolling all eligible clients in community ART distribution points |
| 8 | HP-only | Enrolling all eligible clients in health post |
| 9 | AGs-only | Enrolling all eligible clients in adherence groups |
| *Scenario 10-19: Scenario 5-9: Scenarios for all eligible clients, utilising DSD models with no age/setting specific restrictions, enrolling all clients in two DSD models at a time -* *paired all possible combinations. Clients distribution: 94.7% DSD coverage and 5.3% conventional care not eligible for DSD.* | | |
| 10 | FTRs & 6MMD | Equal distribution of all eligible clients between fast track refills and 6MMD |
| 11 | FTRs & AGs | Equal distribution of all eligible clients between fast track refills and adherence groups |
| 12 | FTRs & CADP | Equal distribution of all eligible clients between fast track refills and community ART distribution points |
| 13 | FTRs & HP | Equal distribution of all eligible clients between fast track refills and health posts |
| 14 | 6MMD & AGs | Equal distribution of all eligible clients between 6MMD and adherence groups |
| 15 | 6MMD & CADP | Equal distribution of all eligible clients between 6MMD and community ART distribution points |
| 16 | 6MMD & HP | Equal distribution of all eligible clients between 6MMD and health posts |
| 17 | AGs & CADP | Equal distribution of all eligible clients between adherence groups and community ART distribution points |
| 18 | AGs & HP | Equal distribution of all eligible clients between adherence groups and health posts |
| 19 | CADP & HP | Equal distribution of all eligible clients between community ART distribution points and health posts |
| *Scenario 20-29: Scenarios for all eligible clients, utilising DSD models with no age/setting specific restrictions, enrolling all clients in three DSD models at a time - paired all possible combinations. Clients distribution: 94.7% DSD coverage and 5.3% conventional care not eligible for DSD.* | | |
| 20 | FTRs, 6MMD & AGs | Equal distribution of all eligible clients between fast-track refills, 6MMD and adherence groups |
| 21 | FTRs, 6MMD & CADP | Equal distribution of all eligible clients between fast-track refills, 6MMD and community ART distribution points |
| 22 | FTRs, 6MMD & HP | Equal distribution of all eligible clients between fast-track refills, 6MMD and health posts |
| 23 | FTRs, AGs & CADP | Equal distribution of all eligible clients between fast-track refills, adherence groups and community ART distribution points |
| 24 | FTR, AGs & HP | Equal distribution of all eligible clients between fast-track refills, adherence groups and health posts |
| 25 | FTRs, CADP & HP | Equal distribution of all eligible clients between fast-track refills, community ART distribution points and health post |
| 26 | 6MMD, AGs & CADP | Equal distribution of all eligible clients between 6MMD, adherence groups and community ART distribution points |
| 27 | 6MMD, AGs & HP | Equal distribution of all eligible clients between 6MMD, adherence groups and health posts |
| 28 | 6MMD, CADP & HP | Equal distribution of all eligible clients between 6MMD, community ART distribution points and health posts |
| 29 | AGs, CADP & HP | Equal distribution of all eligible clients between Adherence groups, community ART distribution points and health posts |
| *Scenario 30-35: Scenarios for all eligible clients, utilising DSD models with no age/setting specific restrictions, enrolling all clients in four DSD models at a time - paired all possible combinations. Clients distribution: 94.7% DSD coverage and 5.3% conventional care not eligible for DSD.* | | |
| 30 | FTRs, MMD6, AGs & CADP | Equal distribution of all eligible clients between fast track refills, 6MMD, adherence groups and community ART distribution points |
| 31 | FTRs, MMD6, AGs & HP | Equal distribution of all eligible clients between fast track refills, 6MMD, adherence groups and health posts |
| 32 | FTRs, MMD6, CADP & HP | Equal distribution of all eligible clients between fast track refills, 6MMD, community ART distribution points and health posts |
| 33 | FTRs, AGs, CADP & HP | Equal distribution of all eligible clients between fast track refills, adherence groups, community ART distribution points and health posts |
| 34 | MMD6, AGs, CADP & HP | Equal distribution of all eligible clients between 6MMD, adherence groups, community ART distribution points and health posts |
|  | ***Scenario 35****: Scenario for all eligible clients, utilising DSD models with no age/setting specific restrictions, enrolling all clients in five DSD models at a time. Clients distribution: 94.7% DSD coverage and 5.3% conventional care not eligible for DSD.* | |
| 35 | FTRs, MMD6, AGs, CADP & HP | Equal distribution of all eligible clients between fast track refills, 6MMD, adherence groups, community ART distribution points and health posts |
| *Scenario 36-42: Age-specific scenarios - paired all possible combinations. Clients distribution: 94.7% DSD coverage (7.1% scholar/adolescent model & 92.9% in each of the DSD model) and 5.3% conventional care not eligible for DSD.* | | |
| 36 | Scholar/adolescent model & FTRs | Enrolling all eligible clients aged ≤24 years in scholar model and clients aged ≥25 years in fast tract refills |
| 37 | Scholar/adolescent model & 6MMD | Enrolling all eligible clients aged ≤24 years in scholar model and clients aged ≥25 years in 6MMD |
| 38 | Scholar/adolescent model & AGs | Enrolling all eligible clients aged ≤24 years in scholar model and clients aged ≥25 years in adherence groups |
| 39 | Scholar/adolescent model & CADP | Enrolling all eligible clients aged ≤24 years in scholar model and all clients aged ≥25 years in community ART distribution points |
| 40 | Scholar/adolescent model & HP | Enrolling all eligible clients aged ≤24 years in scholar model and clients aged ≥25 years in health posts |
| 41 | Scholar/adolescent model & ECH | Enrolling all eligible clients aged ≤24 years in scholar model and clients aged ≥25 years in extended clinic hours |
| 42 | Scholar/adolescent model & HAD | Enrolling all eligible clients aged ≤24 years in scholar model and eligible clients aged ≥25 years in Home ART delivery |
| *Scenario 43-47: scenarios enrolling all eligible clients in a mix of DSD model with age restrictions and models without age restrictions, distributing clients equally across- paired all possible combinations. Clients distribution: 94.7% DSD coverage (3.6% scholar/adolescent model & 96.4% in each DSD model) and 5.3% conventional care not eligible for DSD.* | | |
| 43 | Scholar/adolescent model & fast track refills | Equal distribution of clients aged ≤24 years between scholar/adolescent model and fast track refills and enrolling eligible clients aged ≥25 years in fast track refills |
| 44 | Scholar/adolescent model & 6MMD | Equal distribution of clients aged ≤24 years between scholar/adolescent model and fast track refills and enrolling eligible clients aged ≥25 years in 6MMD |
| 45 | Scholar/adolescent model & AGs | Equal distribution of clients aged ≤24 years between scholar/adolescent model and fast track refills and enrolling eligible clients aged ≥25 years in adherence groups |
| 46 | Scholar/adolescent model & CADP | Equal distribution of clients aged ≤24 years between scholar/adolescent model and fast track refills and enrolling eligible clients aged ≥25 years in community ART distribution points |
| 47 | Scholar/adolescent model & HP | Equal distribution of clients aged ≤24 years between scholar/adolescent model and fast track refills and enrolling eligible clients aged ≥25 years in health posts |
| *Scenario 48-57:* *Age-specific scenarios - paired all possible combinations. Clients distribution: 94.7% DSD coverage (7.1% in each DSD models & 92.9% in either extended clinic hours and Home ART delivery) and 5.3% conventional care not eligible for DSD.* | | |
| 48 | FTRs & ECH | Enrolling all eligible clients aged ≤24 years in fast track refills and clients aged ≥25 years in extended clinic hours |
| 49 | AGs & ECH | Enrolling all eligible clients aged ≤24 years in adherence groups and clients aged ≥25 years in extended clinic hours |
| 50 | CADP & ECH | Enrolling all eligible clients aged ≤24 years in community ART distribution points and clients aged ≥25 years in extended clinic hours |
| 51 | HP & ECH | Enrolling all eligible clients aged ≤24 years in health posts and clients aged ≥25 years in extended clinic hours |
| 52 | 6MMD & ECH | Enrolling all eligible clients aged ≤24 years in 6MMD and clients aged ≥25 years in extended clinic hours |
| 53 | FTRs & HAD | Enrolling all eligible clients aged ≤24 years in fast track refills and clients aged ≥25 years in home ART delivery |
| 54 | CADP & HAD | Enrolling all eligible clients aged ≤24 years in community ART distribution points and clients aged ≥25 years in home ART delivery |
| 55 | AGs & HAD | Enrolling all eligible clients aged ≤24 years in adherence groups and clients aged ≥25 years in home ART delivery |
| 56 | HP & HAD | Enrolling all eligible clients aged ≤24 years in health posts and clients aged ≥25 years in home ART delivery |
| 57 | 6MMD & HAD | Enrolling all eligible clients aged ≤24 years in 6MMD and clients aged ≥25 years in home ART delivery |
| *Scenario 58-87: Age-specific scenarios - paired all possible combinations. Clients distribution: 94.7% DSD coverage (7.1% of clients aged ≤24 in distributed equally in each of the two models & 92.9% of clients in either home ART delivery or extended clinic hours) and 5.3% conventional care not eligible for DSD.* | | |
| 58 | Scholar/adolescent model, FTRs & ECH | Equal distribution of client’s eligible clients aged ≤24 years between scholar/adolescent model and fast track refills and clients aged ≥25 years in extended clinic hours |
| 59 | Scholar/adolescent model, AGs & ECH | Equal distribution of clients aged ≤24 years between scholar/adolescent model and adherence groups and clients aged ≥25 years in extended clinic hours |
| 60 | Scholar/adolescent model, CADP & ECH | Equal distribution of clients aged ≤24 years between scholar/adolescent model and community ART distribution points and clients aged ≥25 years in extended clinic hours |
| 61 | Scholar/adolescent model, 6MMD & ECH | Equal distribution of clients aged ≤24 years between scholar/adolescent model and 6MMD and clients aged ≥25 years in extended clinic hours |
| 62 | Scholar/adolescent model, HP & ECH | Equal distribution of clients aged ≤24 years between scholar/adolescent model and health posts and clients aged ≥25 years in home ART delivery |
| 63 | FTRs, AGs & ECH | Equal distribution of clients aged ≤24 years between fast track refills and adherence groups and clients aged ≥25 years in extended clinic hours |
| 64 | FTRs, CADP & ECH | Equal distribution of clients aged ≤24 years between fast track refills and community adherence groups and clients aged ≥25 years in extended clinic hours |
| 65 | FTRs, 6MMD & ECH | Equal distribution of clients aged ≤24 years between fast track refills and 6MMD and clients aged ≥25 years in extended clinic hours |
| 66 | AGs, CADP & ECH | Equal distribution of clients aged ≤24 years between adherence groups and community adherence groups and clients aged ≥25 years in extended clinic hours |
| 67 | AGS, 6MMD & ECH | Equal distribution of clients aged ≤24 years between adherence groups and 6MMD and clients aged ≥25 years in extended clinic hours |
| 68 | CADP, 6MMD & ECH | Equal distribution of clients aged ≤24 years between community ART distribution points and 6MMD and clients aged ≥25 years in extended clinic hours |
| 69 | Scholar/adolescent model, HP & ECH | Equal distribution of clients aged ≤24 years between scholar/adolescent model and health posts and clients aged ≥25 years in extended clinic hours |
| 70 | FTRs, HP & ECH | Equal distribution of clients aged ≤24 years between scholar/adolescent model and health posts and clients aged ≥25 years in extended clinic hours |
| 71 | AGs, HP & ECH | Equal distribution of clients aged ≤24 years between adherence groups and health posts and clients aged ≥25 years in extended clinic hours |
| 72 | CADP, HP & ECH | Equal distribution of clients aged ≤24 years between community adherence groups and health posts and clients aged ≥25 years in extended clinic hours |
| 73 | 6MMD, HP & ECH | Equal distribution of clients aged ≤24 years between 6MMD and health posts and clients aged ≥25 years in extended clinic hours |
| 73 | Scholar/adolescent model, FTRs & HAD | Equal distribution of clients aged ≤24 years between scholar/adolescent model and fast track refills and clients aged ≥25 years in home ART delivery |
| 74 | Scholar/adolescent model, AGS & HAD | Equal distribution of clients aged ≤24 years between scholar/adolescent model and adherence groups and clients aged ≥25 years in home ART delivery |
| 75 | Scholar/adolescent model, CADP & HAD | Equal distribution of clients aged ≤24 years between scholar/adolescent model and community ART distribution points and clients aged ≥25 years in home ART delivery |
| 76 | Scholar/adolescent model, 6MMD & HAD | Equal distribution of clients aged ≤24 years between scholar/adolescent model and 6MMD and clients aged ≥25 years in home ART delivery |
| 77 | FTRs, AGs & HAD | Equal distribution of clients aged ≤24 years between fast track refills and adherence groups and clients aged ≥25 years in home ART delivery |
| 78 | FTRs, CADP & HAD | Equal distribution of clients aged ≤24 years between fast track refills and community ART distribution points and clients aged ≥25 years in home ART delivery |
| 79 | FTRs, 6MMD & HAD | Equal distribution of clients aged ≤24 years between fast track refills and c and clients aged ≥25 years in home ART delivery |
| 80 | AGs, CADP & HAD | Equal distribution of clients aged ≤24 years between adherence groups and community ART distribution points and clients aged ≥25 years in home ART delivery |
| 81 | AGs, 6MMD & HAD | Equal distribution of clients aged ≤24 years between adherence groups and 6MMD and clients aged ≥25 years in home ART delivery |
| 82 | CADP, 6MMD & HAD | Equal distribution of clients aged ≤24 years between community ART distribution points and 6MMD and clients aged ≥25 years in home ART delivery |
| 83 | Scholar/adolescent model, HP & HAD | Equal distribution of clients aged ≤24 years between scholar/adolescent model and health posts and clients aged ≥25 years in home ART delivery |
| 84 | FTRs, HP & HAD | Equal distribution of clients aged ≤24 years between fast track refills and health posts and clients aged ≥25 years in home ART delivery |
| 85 | AGs, HP& HAD | Equal distribution of clients aged ≤24 years between fast track refills and health posts and clients aged ≥25 years in home ART delivery |
| 86 | CADP, HP & HAD | Equal distribution of clients aged ≤24 years between community ART distribution points and health posts and clients aged ≥25 years in home ART delivery |
| 87 | 6MMD, HP & HAD | Equal distribution of clients aged ≤24 years between 6MMD and health posts and clients aged ≥25 years in home ART delivery |
| *Scenario 88-108: Age-specific scenarios - paired all possible combinations. Clients distribution: 94.7% DSD coverage (7.1% scholar/adolescent model & 92.9% each of the two models) and 5.3% conventional care not eligible for DSD.* | | |
| 88 | Scholar/adolescent model, FTRs & AGs | Clients aged ≤24 years in scholar/adolescent and equal distribution of clients aged ≥25 years in fast track refills and adherence groups. |
| 89 | Scholar/adolescent model, FTRs & CADP | Clients aged ≤24 years in scholar/adolescent and equal distribution of clients aged ≥25 years in fast track refills and community ART distribution points. |
| 90 | Scholar/adolescent model, FTRs & 6MMD | Clients aged ≤24 years in scholar/adolescent and equal distribution of clients aged ≥25 years in fast track refills and 6MMD. |
| 91 | Scholar/adolescent model, FTRs & HP | Clients aged ≤24 years in scholar/adolescent and equal distribution of clients aged ≥25 years in fast track refills and health posts. |
| 92 | Scholar/adolescent model, FTRs & HAD | Clients aged ≤24 years in scholar/adolescent and equal distribution of clients aged ≥25 years in fast track refills and home ART delivery. |
| 93 | Scholar/adolescent model, AGs & CADP | Clients aged ≤24 years in scholar/adolescent and equal distribution of clients aged ≥25 years in adherence groups and community ART distribution points. |
| 94 | Scholar/adolescent model, AGs & 6MMD | Clients aged ≤24 years in scholar/adolescent and equal distribution of clients aged ≥25 years in adherence groups and 6MMD. |
| 95 | Scholar/adolescent model, AGs & HP | Clients aged ≤24 years in scholar/adolescent and equal distribution of clients aged ≥25 years in adherence groups and health posts. |
| 96 | Scholar/adolescent model, AGs & HAD | Clients aged ≤24 years in scholar/adolescent and equal distribution of clients aged ≥25 years in community ART distribution points and home ART delivery. |
| 97 | Scholar/adolescent model, CADP & 6MMD | Clients aged ≤24 years in scholar/adolescent and equal distribution of clients aged ≥25 years in community ART distribution points and 6MMD. |
| 98 | Scholar/adolescent model, CADP & HP | Clients aged ≤24 years in scholar/adolescent and equal distribution of clients aged ≥25 years in community ART distribution points and health posts. |
| 99 | Scholar/adolescent model, CADP & HAD | Clients aged ≤24 years in scholar/adolescent and equal distribution of clients aged ≥25 years in community ART distribution points and home ART delivery. |
| 100 | Scholar/adolescent model, 6MMD & HP | Clients aged ≤24 years in scholar/adolescent and equal distribution of clients aged ≥25 years in 6MMD and health posts. |
| 101 | Scholar/adolescent model, 6MMD & HAD | Clients aged ≤24 years in scholar/adolescent and equal distribution of clients aged ≥25 years in 6MMD and home ART delivery. |
| 102 | Scholar/adolescent model, HP & HAD | Clients aged ≤24 years in scholar/adolescent and equal distribution of clients aged ≥25 years in health posts and home ART delivery. |
| 103 | Scholar/adolescent model, FTRs & ECH | Clients aged ≤24 years in scholar/adolescent and equal distribution of clients aged ≥25 years in fast track refills and extended clinic hours. |
| 104 | Scholar/adolescent model, AGs & ECH | Clients aged ≤24 years in scholar/adolescent and equal distribution of clients aged ≥25 years in adherence groups and extended clinic hours. |
| 105 | Scholar/adolescent model, CADP & ECH | Clients aged ≤24 years in scholar/adolescent and equal distribution of clients aged ≥25 years in community ART distribution points and extended clinic hours. |
| 106 | Scholar/adolescent model, 6MMD & ECH | Clients aged ≤24 years in scholar/adolescent and equal distribution of clients aged ≥25 years in 6MMD and extended clinic hours. |
| 107 | Scholar/adolescent model, HP & ECH | Clients aged ≤24 years in scholar/adolescent and equal distribution of clients aged ≥25 years in health posts and extended clinic hours. |
| 108 | Scholar/adolescent model, HAD & ECH | Clients aged ≤24 years in scholar/adolescent and equal distribution of clients aged ≥25 years in home ART delivery and extended clinic hours. |
| *Scenario 109-118: Settings-specific scenarios - paired all possible combinations. Clients distribution: 94.7% DSD coverage and 5.3% conventional care not eligible for DSD (30.9% mobile ART delivery & 69.1% in each of DSD models)* | | |
| 109 | MAD & FTRs | Equal distribution of clients in rural area between mobile ART delivery and fast track refills and clients in urban areas only in fast track refills. |
| 110 | MAD & MMD6 | Equal distribution of clients in rural area between mobile ART delivery and 6MMD and clients in urban areas only in 6MMD. |
| 111 | MAD & AGs | Equal distribution of clients in rural area between mobile ART delivery and adherence groups and clients in urban areas only in adherence groups. |
| 112 | MAD & CADP | Equal distribution of clients in rural area between mobile ART delivery and community ART distribution points and clients in urban areas only in community ART distribution points. |
| 113 | MAD & HP | Equal distribution of clients in rural area between mobile ART delivery and health posts and clients in urban areas only in health posts. |
| 114 | MAD & FTRs | Enrolling all eligible clients in rural areas in mobile ART delivery and clients in urban areas in fast track refills. |
| 115 | MAD & 6MMD | Enrolling all eligible clients in rural areas in mobile ART delivery and clients in urban areas in 6MMD. |
| 116 | MAD & AGs | Enrolling all eligible clients in rural areas in mobile ART delivery and clients in urban areas in adherence groups. |
| 117 | MAD & CADP | Enrolling all eligible clients in rural areas in mobile ART delivery and clients in urban areas in community ART distribution points. |
| 118 | MAD & HP | Enrolling all eligible clients in rural areas in mobile ART delivery and clients in urban areas in health posts. |
| *Scenario 119-125: Settings and age specific scenarios - paired all possible combinations. Clients distribution: 94.7% DSD coverage and 5.3% conventional care not eligible for DSD (4.8% scholar/adolescent model, 64.3% each of the DSD model & 30.9% mobile ART delivery).* | | |
| 119 | Scholar/adolescent model, MAD & FTRs | Enrolling all eligible clients aged ≤24 in urban areas in scholar/adolescent model and clients all clients in rural areas in mobile ART delivery and clients aged ≥25 years in urban settings in fast track refills. |
| 120 | Scholar/adolescent model, MAD & 6MMD | Enrolling all eligible clients aged ≤24 in urban areas in scholar/adolescent model and clients all clients in rural areas in mobile ART delivery and clients aged ≥25 years in urban settings in 6MMD. |
| 121 | Scholar/adolescent model, MAD & AGs | Enrolling all eligible clients aged ≤24 in urban areas in scholar/adolescent model and clients all clients in rural areas in mobile ART delivery and clients aged ≥25 years in urban settings in adherence groups. |
| 122 | Scholar/adolescent model, MAD & CADP | Enrolling all eligible clients aged ≤24 in urban areas in scholar/adolescent model and clients all clients in rural areas in mobile ART delivery and clients aged ≥25 years in urban settings in community ART distribution points. |
| 123 | Scholar/adolescent model, MAD & HP | Enrolling all eligible clients aged ≤24 in urban areas in scholar/adolescent model and clients all clients in rural areas in mobile ART delivery and clients aged ≥25 years in urban settings in health posts. |
| 124 | Scholar/adolescent model, MAD & ECH | Enrolling all eligible clients aged ≤24 in urban areas in scholar/adolescent model and clients all clients in rural areas in mobile ART delivery and clients aged ≥25 years in urban settings in extended clinic hours. |
| 125 | Scholar/adolescent model, MAD & HAD | Enrolling all eligible clients aged ≤24 in urban areas in scholar/adolescent model and clients all clients in rural areas in mobile ART delivery and clients aged ≥25 years in urban settings in Home ART delivery. |

### Table S5. Cost-effectiveness analysis of all scenarios

| **Scenario** | **Scenario name** | **Total number retained** | **Total number suppressed** | **Total Provider costs** | **Total cost to Clients** | **ICER** |
| --- | --- | --- | --- | --- | --- | --- |
| Base case | Current distribution | 817,948 | 770,086 | $84,332,234 | $4,145,400 | Strongly dominated |
| 1 | Scholar/adolescent model-only | 798,346 | 736,820 | $87,280,581 | $5,581,838 | Strongly dominated |
| 2 | MAD-only | 813,319 | 744,752 | $93,139,757 | $4,743,071 | Strongly dominated |
| 3 | HAD-only | 837,820 | 792,967 | $109,110,439 | $5,050,269 | Strongly dominated |
| 4 | ECH-only | 802,451 | 774,257 | $87,728,203 | $2,891,995 | Strongly dominated |
| 5 | FTRs-only | 840,869 | 804,632 | $89,388,584 | $5,398,622 | $482 |
| 6 | 6MMD-only | 827,415 | 782,545 | $83,095,136 | $3,455,290 | Cost-saving (*compared to base case)* |
| 7 | CADP-only | 838,325 | 798,779 | $89,350,068 | $5,354,108 | Strongly dominated |
| 8 | HP-only | 813,234 | 772,556 | $87,524,461 | $5,195,471 | Strongly dominated |
| 9 | AGs-only | 835,581 | 800,878 | $87,577,961 | $10,056,934 | $245 |
| 10 | FTRs & 6MMD | 834,142 | 793,588 | $86,241,860 | $4,426,956 | Weakly dominated |
| 11 | FTRs & AGs | 838,225 | 802,755 | $88,483,273 | $7,727,778 | $482 |
| 12 | FTRs & CADP | 839,597 | 801,705 | $89,369,326 | $5,376,365 | Strongly dominated |
| 13 | FTRs & HP | 827,052 | 788,594 | $88,456,522 | $5,297,046 | Strongly dominated |
| 14 | 6MMD & AGs | 831,498 | 791,712 | $85,336,549 | $6,756,112 | $245 |
| 15 | 6MMD & CADP | 832,870 | 790,662 | $86,222,602 | $4,404,699 | Strongly dominated |
| 16 | 6MMD & HP | 820,325 | 777,551 | $85,309,798 | $4,325,380 | Strongly dominated |
| 17 | AGs & CADP | 836,953 | 799,829 | $88,464,015 | $7,705,521 | Strongly dominated |
| 18 | AGs & HP | 824,408 | 786,717 | $87,551,211 | $7,626,202 | Strongly dominated |
| 19 | CADP & HP | 825,779 | 785,668 | $88,437,264 | $5,274,789 | Strongly dominated |
| 20 | FTRs, 6MMD & AGs | 834,622 | 796,018 | $86,687,227 | $6,303,615 | Weakly dominated |
| 21 | FTRs, 6MMD & CADP | 835,536 | 795,319 | $87,277,929 | $4,736,006 | Strongly dominated |
| 22 | FTRs, 6MMD & HP | 827,173 | 786,578 | $86,669,394 | $4,683,127 | Strongly dominated |
| 23 | FTRs, AGs & CADP | 838,258 | 801,430 | $88,772,204 | $6,936,554 | Strongly dominated |
| 24 | FTR, AGs & HP | 829,895 | 792,689 | $88,163,669 | $6,883,675 | Strongly dominated |
| 25 | FTRs, CADP & HP | 830,809 | 791,989 | $88,754,371 | $5,316,067 | Strongly dominated |
| 26 | 6MMD, AGs & CADP | 833,774 | 794,068 | $86,674,388 | $6,288,777 | Weakly dominated |
| 27 | 6MMD, AGs & HP | 825,410 | 785,327 | $86,065,853 | $6,235,898 | Strongly dominated |
| 28 | 6MMD, CADP & HP | 826,325 | 784,627 | $86,656,555 | $4,668,290 | Strongly dominated |
| 29 | AGs, CADP & HP | 829,047 | 790,738 | $88,150,830 | $6,868,838 | Strongly dominated |
| 30 | FTRs, MMD6, AGs & CADP | 835,548 | 796,709 | $87,352,937 | $6,066,238 | Weakly dominated |
| 31 | FTRs, MMD6, AGs & HP | 829,275 | 790,153 | $86,896,535 | $6,026,579 | Strongly dominated |
| 32 | FTRs, MMD6, CADP & HP | 829,961 | 789,628 | $87,339,562 | $4,850,873 | Strongly dominated |
| 33 | FTRs, AGs, CADP & HP | 832,002 | 794,211 | $88,460,268 | $6,501,284 | Strongly dominated |
| 34 | MMD6, AGs, CADP & HP | 828,639 | 788,690 | $86,886,906 | $6,015,451 | Strongly dominated |
| 35 | FTRs, MMD6, AGs, CADP & HP | 831,085 | 791,878 | $87,387,242 | $5,892,085 | Strongly dominated |
| 36 | Scholar/adolescent model & FTRs | 844,148 | 801,920 | $89,919,547 | $5,709,962 | Strongly dominated |
| 37 | Scholar/adolescent model & 6MMD | 830,515 | 783,863 | $83,947,939 | $3,895,498 | Weakly dominated |
| 38 | Scholar/adolescent model & AGs | 835,293 | 794,629 | $87,823,118 | $10,000,680 | Strongly dominated |
| 39 | Scholar/adolescent model & CADP | 840,242 | 803,475 | $89,720,660 | $5,658,795 | Strongly dominated |
| 40 | Scholar/adolescent model & HP | 816,012 | 769,660 | $87,926,246 | $5,505,595 | Strongly dominated |
| 41 | Scholar/adolescent model & ECH | 808,952 | 782,397 | $88,437,136 | $3,211,782 | Strongly dominated |
| 42 | Scholar/adolescent model & HAD | 844,321 | 801,106 | $109,819,372 | $5,370,056 | Strongly dominated |
| 43 | Scholar/adolescent model & fast track refills | 842,508 | 803,276 | $89,654,065 | $5,554,292 | Strongly dominated |
| 44 | Scholar/adolescent model & 6MMD | 828,965 | 783,204 | $83,521,537 | $3,675,394 | Weakly dominated |
| 45 | Scholar/adolescent model & AGs | 835,437 | 797,753 | $87,700,540 | $10,028,807 | Strongly dominated |
| 46 | Scholar/adolescent model & CADP | 839,283 | 801,127 | $89,535,364 | $5,506,451 | Strongly dominated |
| 47 | Scholar/adolescent model & HP | 814,623 | 771,108 | $87,725,354 | $5,350,533 | Strongly dominated |
| 48 | FTRs & ECH | 805,673 | 785,109 | $87,906,174 | $2,900,441 | Strongly dominated |
| 49 | AGs & ECH | 809,240 | 788,646 | $88,191,979 | $3,268,035 | Strongly dominated |
| 50 | CADP & ECH | 807,034 | 777,701 | $88,066,544 | $2,907,095 | Strongly dominated |
| 51 | HP & ECH | 806,174 | 785,293 | $88,035,351 | $2,901,658 | Strongly dominated |
| 52 | 6MMD & ECH | 805,851 | 781,080 | $87,584,333 | $2,771,574 | Strongly dominated |
| 53 | FTRs & HAD | 841,042 | 803,818 | $109,288,410 | $5,058,716 | Strongly dominated |
| 54 | CADP & HAD | 841,220 | 799,789 | $108,966,569 | $4,929,848 | Strongly dominated |
| 55 | AGs & HAD | 844,609 | 807,356 | $109,574,215 | $5,426,310 | $7,409 |
| 56 | HP & HAD | 842,403 | 796,410 | $109,448,780 | $5,065,369 | Strongly dominated |
| 57 | 6MMD & HAD | 841,543 | 804,002 | $109,417,587 | $5,059,933 | Strongly dominated |
| 58 | Scholar/adolescent model, FTRs & ECH | 807,313 | 783,753 | $88,171,655 | $3,056,111 | Strongly dominated |
| 59 | Scholar/adolescent model, AGs & ECH | 809,096 | 785,522 | $88,314,558 | $3,239,909 | Strongly dominated |
| 60 | Scholar/adolescent model, CADP & ECH | 807,993 | 780,049 | $88,251,840 | $3,059,438 | Strongly dominated |
| 61 | Scholar/adolescent model, 6MMD & ECH | 807,401 | 781,738 | $88,010,735 | $2,991,678 | Strongly dominated |
| 62 | Scholar/adolescent model, HP & ECH | 807,457 | 786,878 | $88,049,076 | $3,084,238 | Strongly dominated |
| 63 | FTRs, AGs & ECH | 806,354 | 781,405 | $87,986,359 | $2,903,768 | Strongly dominated |
| 64 | FTRs, CADP & ECH | 805,762 | 783,094 | $87,745,254 | $2,836,008 | Strongly dominated |
| 65 | FTRs, 6MMD & ECH | 808,137 | 783,174 | $88,129,262 | $3,087,565 | Strongly dominated |
| 66 | AGs, CADP & ECH | 807,546 | 784,863 | $87,888,156 | $3,019,805 | Strongly dominated |
| 67 | AGS, 6MMD & ECH | 806,443 | 779,390 | $87,825,439 | $2,839,334 | Strongly dominated |
| 68 | CADP, 6MMD & ECH | 807,563 | 783,845 | $88,236,244 | $3,056,720 | Strongly dominated |
| 69 | Scholar/adolescent model, HP & ECH | 805,924 | 785,201 | $87,970,762 | $2,901,050 | Strongly dominated |
| 70 | FTRs, HP & ECH | 807,707 | 786,970 | $88,113,665 | $3,084,847 | Strongly dominated |
| 71 | AGs, HP & ECH | 806,604 | 781,497 | $88,050,947 | $2,904,376 | Strongly dominated |
| 72 | CADP, HP & ECH | 806,013 | 783,186 | $87,809,842 | $2,836,616 | Strongly dominated |
| 73 | 6MMD, HP & ECH | 842,681 | 802,462 | $109,553,891 | $5,214,386 | Strongly dominated |
| 74 | Scholar/adolescent model, FTRs & HAD | 844,465 | 804,231 | $109,696,793 | $5,398,183 | Strongly dominated |
| 75 | Scholar/adolescent model, AGS & HAD | 843,362 | 798,758 | $109,634,076 | $5,217,713 | Strongly dominated |
| 76 | Scholar/adolescent model, CADP & HAD | 842,770 | 800,448 | $109,392,971 | $5,149,952 | Strongly dominated |
| 77 | Scholar/adolescent model, 6MMD & HAD | 842,826 | 805,587 | $109,431,312 | $5,242,513 | Weakly dominated |
| 78 | FTRs, AGs & HAD | 841,722 | 800,114 | $109,368,595 | $5,062,042 | Strongly dominated |
| 79 | FTRs, CADP & HAD | 841,131 | 801,804 | $109,127,489 | $4,994,282 | Strongly dominated |
| 80 | FTRs, 6MMD & HAD | 843,506 | 801,883 | $109,511,497 | $5,245,840 | Strongly dominated |
| 81 | AGs, CADP & HAD | 842,915 | 803,573 | $109,270,392 | $5,178,079 | Strongly dominated |
| 82 | AGs, 6MMD & HAD | 841,811 | 798,100 | $109,207,675 | $4,997,609 | Strongly dominated |
| 83 | CADP, 6MMD & HAD | 842,932 | 802,554 | $109,618,479 | $5,214,995 | Strongly dominated |
| 84 | Scholar/adolescent model, HP & HAD | 841,293 | 803,910 | $109,352,998 | $5,059,324 | Strongly dominated |
| 85 | FTRs, HP & HAD | 843,076 | 805,679 | $109,495,901 | $5,243,121 | Strongly dominated |
| 86 | AGs, HP& HAD | 841,973 | 800,206 | $109,433,183 | $5,062,651 | Strongly dominated |
| 87 | CADP, HP & HAD | 841,382 | 801,896 | $109,192,078 | $4,994,891 | Strongly dominated |
| 88 | Scholar/adolescent model, FTRs & AGs | 839,720 | 798,274 | $88,871,333 | $7,855,321 | Strongly dominated |
| 89 | Scholar/adolescent model, FTRs & CADP | 842,195 | 802,698 | $89,820,103 | $5,684,379 | Strongly dominated |
| 90 | Scholar/adolescent model, FTRs & 6MMD | 837,332 | 792,891 | $86,933,743 | $4,802,730 | Strongly dominated |
| 91 | Scholar/adolescent model, FTRs & HP | 830,080 | 785,790 | $88,922,896 | $5,607,778 | Strongly dominated |
| 92 | Scholar/adolescent model, FTRs & HAD | 844,234 | 801,513 | $99,869,459 | $5,540,009 | Strongly dominated |
| 93 | Scholar/adolescent model, AGs & CADP | 837,767 | 799,052 | $88,771,889 | $7,829,738 | Strongly dominated |
| 94 | Scholar/adolescent model, AGs & 6MMD | 832,904 | 789,246 | $85,885,529 | $6,948,089 | Strongly dominated |
| 95 | Scholar/adolescent model, AGs & HP | 825,652 | 782,144 | $87,874,682 | $7,753,137 | Strongly dominated |
| 96 | Scholar/adolescent model, AGs & HAD | 839,807 | 797,868 | $98,821,245 | $7,685,368 | Strongly dominated |
| 97 | Scholar/adolescent model, CADP & 6MMD | 835,379 | 793,669 | $86,834,300 | $4,777,146 | Strongly dominated |
| 98 | Scholar/adolescent model, CADP & HP | 828,127 | 786,568 | $88,823,453 | $5,582,195 | Strongly dominated |
| 99 | Scholar/adolescent model, CADP & HAD | 842,281 | 802,291 | $99,770,016 | $5,514,426 | Strongly dominated |
| 100 | Scholar/adolescent model, 6MMD & HP | 823,264 | 776,761 | $85,937,093 | $4,700,546 | Strongly dominated |
| 101 | Scholar/adolescent model, 6MMD & HAD | 837,418 | 792,485 | $96,883,656 | $4,632,777 | Strongly dominated |
| 102 | Scholar/adolescent model, HP & HAD | 830,166 | 785,383 | $98,872,809 | $5,437,825 | Strongly dominated |
| 103 | Scholar/adolescent model, FTRs & ECH | 826,550 | 792,158 | $89,178,341 | $4,460,872 | Strongly dominated |
| 104 | Scholar/adolescent model, AGs & ECH | 822,122 | 788,513 | $88,130,127 | $6,606,231 | Strongly dominated |
| 105 | Scholar/adolescent model, CADP & ECH | 824,597 | 792,936 | $89,078,898 | $4,435,288 | Strongly dominated |
| 106 | Scholar/adolescent model, 6MMD & ECH | 819,734 | 783,130 | $86,192,538 | $3,553,640 | Strongly dominated |
| 107 | Scholar/adolescent model, HP & ECH | 812,482 | 776,029 | $88,181,691 | $4,358,688 | Strongly dominated |
| 108 | Scholar/adolescent model, HAD & ECH | 826,636 | 791,752 | $99,128,254 | $4,290,919 | Strongly dominated |
| 109 | MAD & FTRs | 844,409 | 802,197 | $92,280,254 | $5,120,643 | Strongly dominated |
| 110 | MAD & MMD6 | 831,633 | 780,024 | $86,822,887 | $3,473,242 | Strongly dominated |
| 111 | MAD & AGs | 840,344 | 798,362 | $90,792,565 | $9,060,995 | Strongly dominated |
| 112 | MAD & CADP | 842,674 | 796,220 | $92,291,788 | $5,085,661 | Strongly dominated |
| 113 | MAD & HP | 818,973 | 769,095 | $90,481,332 | $4,935,811 | Strongly dominated |
| 114 | MAD & FTRs | 847,949 | 799,763 | $95,171,925 | $4,842,665 | Strongly dominated |
| 115 | MAD & 6MMD | 835,851 | 777,502 | $90,550,639 | $3,491,194 | Strongly dominated |
| 116 | MAD & AGs | 845,106 | 795,845 | $94,007,169 | $8,065,055 | Strongly dominated |
| 117 | MAD & CADP | 847,024 | 793,661 | $95,233,508 | $4,817,215 | Strongly dominated |
| 118 | MAD & HP | 824,713 | 765,634 | $93,438,203 | $4,676,150 | Strongly dominated |
| 119 | Scholar/adolescent model, MAD & FTRs | 849,742 | 796,324 | $95,484,499 | $5,048,018 | Strongly dominated |
| 120 | Scholar/adolescent model, MAD & 6MMD | 838,126 | 777,829 | $91,140,416 | $3,785,923 | Strongly dominated |
| 121 | Scholar/adolescent model, MAD & AGs | 845,007 | 790,668 | $94,180,566 | $8,028,637 | Strongly dominated |
| 122 | Scholar/adolescent model, MAD & CADP | 848,365 | 795,783 | $95,487,386 | $5,021,027 | Strongly dominated |
| 123 | Scholar/adolescent model, MAD & HP | 827,013 | 763,581 | $93,754,323 | $4,886,029 | Strongly dominated |
| 124 | Scholar/adolescent model, MAD & ECH | 826,467 | 781,074 | $94,573,568 | $3,325,743 | Strongly dominated |
| 125 | Scholar/adolescent model, MAD & HAD | 848,478 | 798,981 | $109,046,436 | $4,805,072 | Strongly dominated |
| 6MMD, six months dispensing; MAD, Mobile ART delivery, HAD, Home ART delivery, ECH, extended clinic hours, FTRs, fast track refills; CADP, community ART distribution points, HP, health posts, AGs, adherence groups | | | | | | |
